# Neutralizing Antibody Response to Pseudotype SARS-CoV-2 Differs between mRNA-1273 and BNT162b2 COVID-19 Vaccines and by History of SARS-CoV-2 Infection

**DOI:** 10.1101/2021.10.20.21265171

**Authors:** Harmony L. Tyner, Jefferey L Burgess, Lauren Grant, Manjusha Gaglani, Jennifer L. Kuntz, Allison L. Naleway, Natalie J. Thornburg, Alberto J. Caban-Martinez, Sarang K. Yoon, Meghan K. Herring, Shawn C. Beitel, Lenee Blanton, Janko Nikolich-Zugich, Matthew S. Thiese, Jessica Flores Pleasants, Ashley L. Fowlkes, Karen Lutrick, Kayan Dunnigan, Young M.Yoo, Spencer Rose, Holly Groom, Jennifer Meece, Meredith G. Wesley, Natasha Schaefer-Solle, Paola Louzado-Feliciano, Laura J. Edwards, Lauren E. W. Olsho, Mark G. Thompson

## Abstract

**Background:** Data on the development of neutralizing antibodies against SARS-CoV-2 after SARS-CoV-2 infection and after vaccination with messenger RNA (mRNA) COVID-19 vaccines are limited.

**Methods:** From a prospective cohort of 3,975 adult essential and frontline workers tested weekly from August, 2020 to March, 2021 for SARS-CoV-2 infection by Reverse Transcription-Polymerase Chain Reaction (RT-PCR) assay irrespective of symptoms, 497 participants had sera drawn after infection (170), vaccination (327), and after both infection and vaccination (50 from the infection population). Serum was collected after infection and each vaccine dose. Serum-neutralizing antibody titers against USA-WA1/2020-spike pseudotype virus were determined by the 50% inhibitory dilution. Geometric mean titers (GMTs) and corresponding fold increases were calculated using t-tests and linear mixed effects models.

**Results:** Among 170 unvaccinated participants with SARS-CoV-2 infection, 158 (93%) developed neutralizing antibodies (nAb) with a GMT of 1,003 (95% CI=766-1,315). Among 139 previously uninfected participants, 138 (99%) developed nAb after mRNA vaccine dose-2 with a GMT of 3,257 (95% CI = 2,596-4,052). GMT was higher among those receiving mRNA-1273 vaccine (GMT =4,698, 95%CI= 3,186-6,926) compared to BNT162b2 vaccine (GMT=2,309, 95%CI=1,825-2,919). Among 32 participants with prior SARS-CoV-2 infection, GMT was 21,655 (95%CI=14,766-31,756) after mRNA vaccine dose-1, without further increase after dose- 2.

**Conclusions:** A single dose of mRNA vaccine after SARS-CoV-2 infection resulted in the highest observed nAb response. Two doses of mRNA vaccine in previously uninfected participants resulted in higher nAb to SARS-CoV-2 than after one dose of vaccine or SARS-CoV-2 infection alone. Neutralizing antibody response also differed by mRNA vaccine product.

**Main Point Summary:** One dose of mRNA COVID-19 vaccine after previous SARS-CoV-2 infection produced the highest neutralizing antibody titers; among those without history of infection, two doses of mRNA vaccine produced the most robust response.

## INTRODUCTION

A humoral immune response to Severe Acute Respiratory Syndrome Coronavirus 2 (SARS-CoV-2) infection or vaccination against COVID-19 is manifested in the development of neutralizing antibodies (nAb) against SARS-CoV-2, the causative agent for coronavirus disease 2019 (COVID-19) and is thought to be predictive of protection against subsequent symptomatic and severe illness [1, 2]. Most studies of nAb response following SARS-CoV-2 infection have been conducted among patients with medically attended and severe symptomatic COVID-19 [3-9]. Less is known about nAb response following mildly symptomatic or asymptomatic infection or following vaccination administered in real-world conditions.

In a multi-center prospective cohort of essential and frontline workers, sera were collected before and after SARS-CoV-2 infection and before and after vaccination with messenger RNA (mRNA) COVID-19 vaccines. This study has four objectives: first, to describe nAb response following SARS-CoV-2 infection and examine whether nAb responses differed by illness characteristics, sociodemographic and health characteristics, viral shedding duration, or peak RNA load, second, to describe nAb responses in previously uninfected participants 2-3 weeks following one or two doses of mRNA vaccines and examine whether nAb response differed by socio-demographic or health characteristics, febrile symptoms, or the use of analgesics before or after vaccination [10]. Response to mRNA-1273 (Moderna) versus BNT162b2 (Pfizer-BioNTech) mRNA vaccines was also compared. A third objective was to describe nAb response following mRNA vaccination among participants who were infected with SARS-CoV-2 prior to vaccination, and fourth, to compare the magnitude of nAb GMT following infection only, vaccination only, and vaccination after SARS-CoV-2 infection.

## METHODS

### Study Design and Setting

HEROES-RECOVER is a network of prospective cohorts of Health Care Personnel, first responders, and essential and frontline workers in eight locations across the United States (Phoenix, Tucson, and other areas in Arizona; Miami, Florida; Duluth, Minnesota; Portland, Oregon; Temple, Texas; and Salt Lake City, Utah) that share a common protocol and methods [11-13]. The cohort of 3,975 participants from which the three analytic samples are drawn has been described in a previous publication [12]. At the time of enrollment, all participants provided written consent. This study was reviewed and approved by Institutional Review Boards (IRBs) at each of the participating institutions. ^§^

### Specimen Collection and Participant-Reported Characteristics and Outcome Measures from August 2020 to March 2021

Participants completed electronic surveys at the time of enrollment that collected sociodemographic and health characteristics. Participants self-collected a mid-turbinate nasal swab weekly and collected an additional mid-turbinate swab and saliva specimen at the onset of COVID-19-like illness (CLI), defined as one or more symptom(s) of fever, chills, cough, shortness of breath, sore throat, diarrhea, muscle aches, and/or a change in smell or taste. When CLI was identified, participants completed electronic surveys at the beginning and end of symptoms to describe duration of illness and the presence or absence of fever, feverishness, chills, or a measured temperature higher than 38 degrees F (100 degrees F).

Participants provided a serum specimen upon enrollment and every 3 months thereafter. Convalescent serum was collected about 30 days after the first SARS-CoV-2 positive test result. Participants who were vaccinated with mRNA vaccines provided a serum specimen approximately 14–21 days after each dose of vaccine.

### Vaccination Status and Patient-Reported Events Following Vaccination

COVID-19 vaccination status was collected through (1) participant self-report by electronic and telephone surveys, (2) direct upload of vaccine card images after each vaccine dose, (3) review of vaccination records in electronic medical records or occupational health records, and (4) review of state immunization information systems in Minnesota, Oregon, Texas, and Utah. Participants reported any fever and use of analgesics before and after each vaccine dose through electronic surveys [12].

### Laboratory Methods

Self-collected mid-turbinate swab specimens were shipped on cold packs and tested for SARS-CoV-2 by qualitative reverse-transcription polymerase chain reaction (RT-PCR) assay at Marshfield Clinic Laboratory (Marshfield, WI). Quantitative RT-PCR was done at the Wisconsin State Laboratory of Hygiene (Madison, Wisconsin). Both tests used methods described in a previous publication [12].

Serum-neutralizing antibodies were measured against wild or United States’ origin USA- WA1/2020-spike pseudotype virus, similar to other evaluations prior to the widespread circulation of SARS-CoV-2 variants of concern [5, 8]. The USA-WA1/2020-spike pseudotype virus is an artificially created virus displaying the spike protein sequenced from a clinical SARS-CoV-2 isolate, providing a standardized viral proxy for response to antibody response. The resulting serum-neutralizing antibodies were calculated as the 50% inhibitory dilution (ID50). These serologic assays, which quantify the amount of host serum needed to neutralize pseudovirus infectivity, an *in-vitro* proxy for antibody mediated protection against SARS-CoV-2, were conducted by LabCorp (South San Francisco, CA) staff blinded to SARS-CoV-2 infection status and vaccine product (Supplementary Appendix: Methods)[14].

### Statistical Analysis

Neutralizing antibodies were analyzed as log10 titers and back transformed to estimate geometric mean titers GMTs. A titer of <40 (negative) was imputed as 20 (half the detectable limit) for analysis. For nAb following infection, 95% confidence intervals (CI) of GMT were calculated using one-sample t-tests. For nAb following vaccination, GMT and 95% CI were calculated using a linear mixed effects model, with a repeated measure for nAb following vaccine dose-1 and dose-2. We evaluated associations between sociodemographic or health characteristics and nAb measures using Pearson’s chi-squared, Fisher’s exact test, or one-way analysis of variance. We considered p-values < .05 as statistically significant. Estimates were considered statistically different if their 95% CI did not overlap. The inclusion of days from infection or vaccination to sera collection and sociodemographic and health characteristics in the adjusted GMT models did not change point estimates by >5% and, thus, were not included in the adjusted models (Supplementary Appendix: Methods).

## RESULTS

### Participant Groups and Characteristics

Analyses included (1) 170 participants with evidence of SARS-CoV-2 infection and post-infection sera, prior to receipt of mRNA-COVID-19 vaccine, (2) 139 participants with no history of SARS-CoV-2 infection, who received mRNA-COVID-19 vaccine and submitted post- vaccination sera, and (3) 50 participants from the group of previously infected participants with post-infection sera, who subsequently received mRNA-COVID-19 vaccine and had post-vaccination sera collected following infection (Figure 1). Of the 50 participants with post-vaccination sera following infection, 42 had sera drawn after dose 1, 40 had sera drawn after dose 2, 32 had sera drawn after both doses (Figure S3). (Supplementary Appendix: Methods: Study Population).

**Figure 1.**
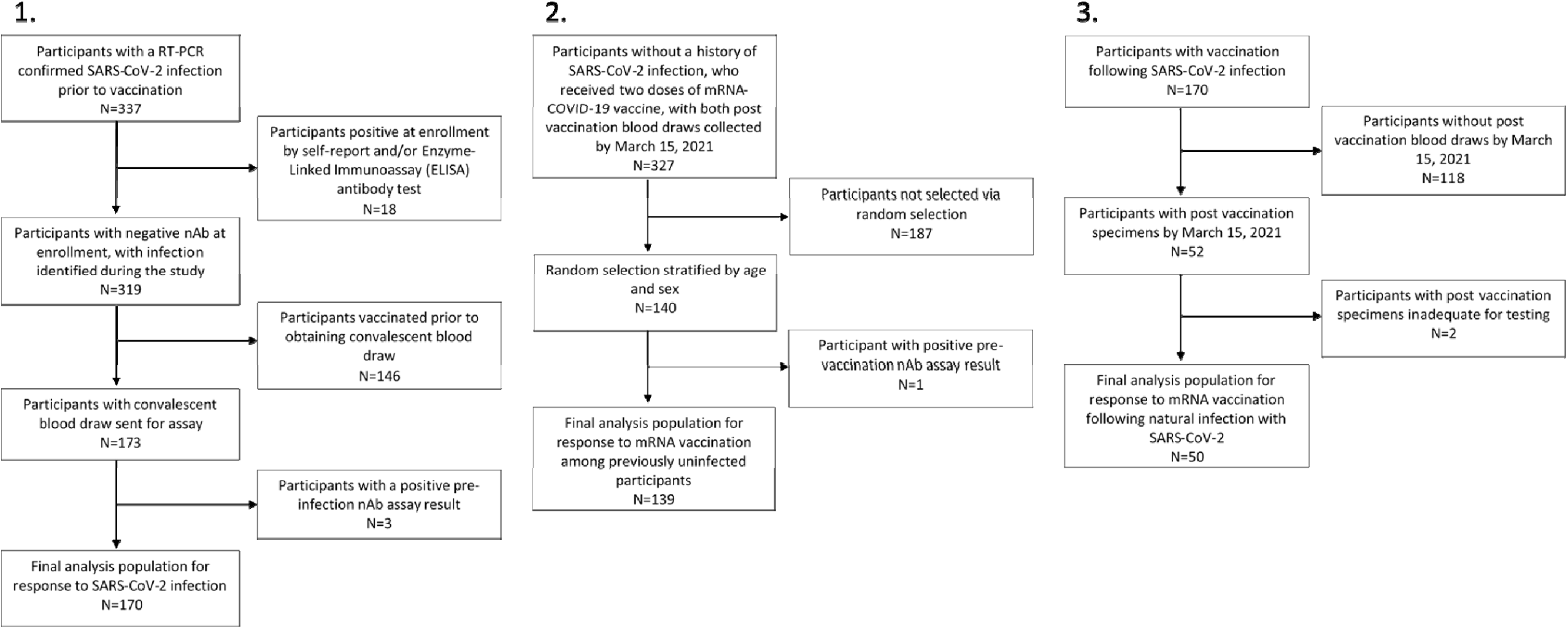
Participant selection for analytic populations. Legend: Panel 1: Selection of analytic population of participants with positive SARS-CoV-2 test results. Panel 2: Selection of analytic population of previously uninfected participants receiving both doses of mRNA vaccine. Panel 3: Selection of analytic population of participants with prior positive SARS-CoV-2 test result receiving both doses of mRNA vaccine.

Sociodemographic and health characteristics were similar across groups at enrollment (Table 1). At least half were female (range across samples = 50%-58%), most were aged 18-49 years (64%-71%), and the majority self-reported their race as White (91%-92%). Occupation was similar across groups; 12-28% were primary health care providers, 39-58% were nurses or other allied health care providers, 19-39% were first responders, and 1-9% were essential and other frontline workers. Most (67%-72%) participants were overweight (BMI 25-29.9 kg/m^2^) or obese (BMI 30+ kg/m^2^). Approximately one-half of participants described themselves as being in excellent or very good health (55%-71%), 22%-30% reported at least one chronic condition and at least one daily medication, and 15%-22% reported currently smoking tobacco products.

**Table 1.**
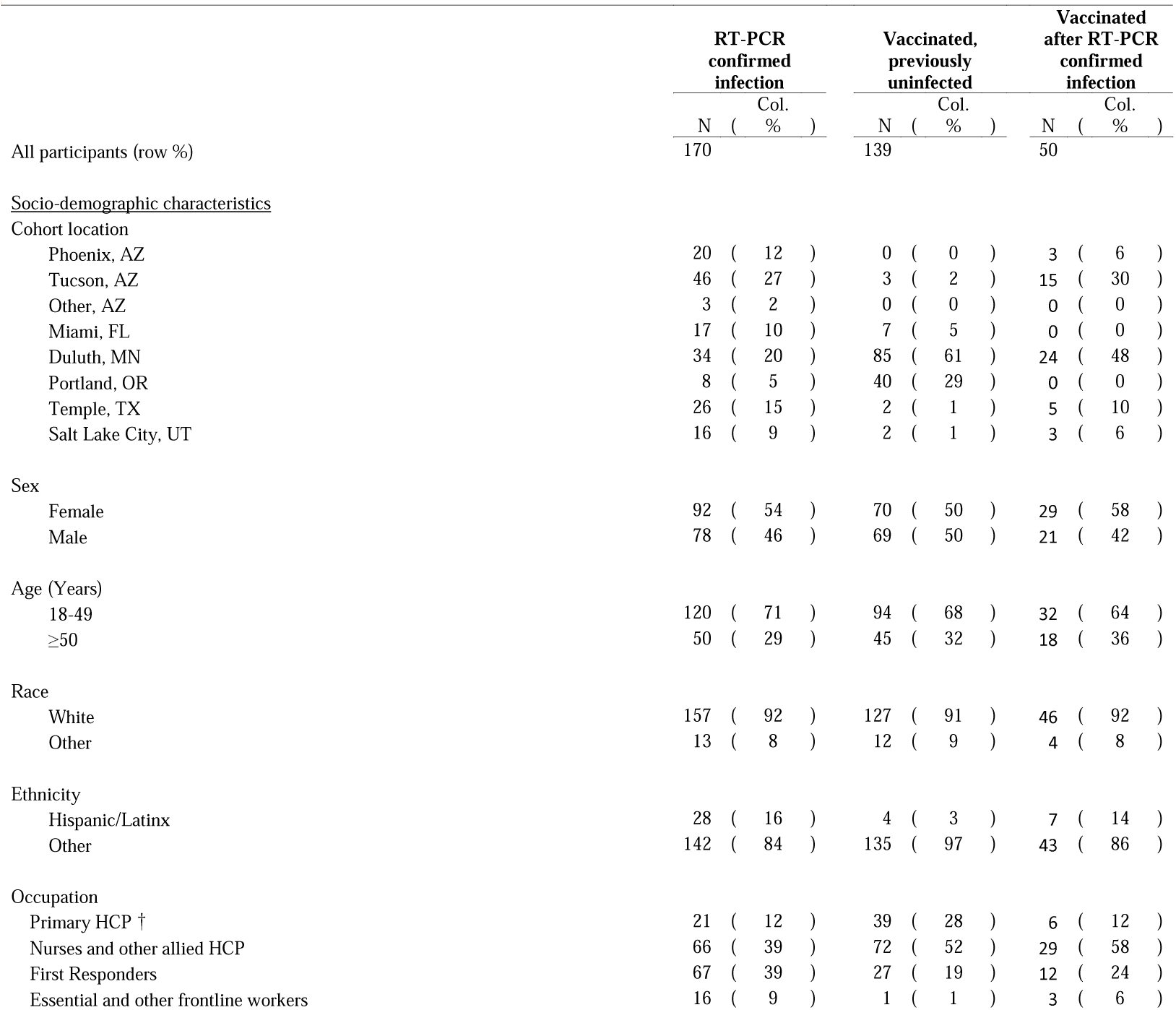
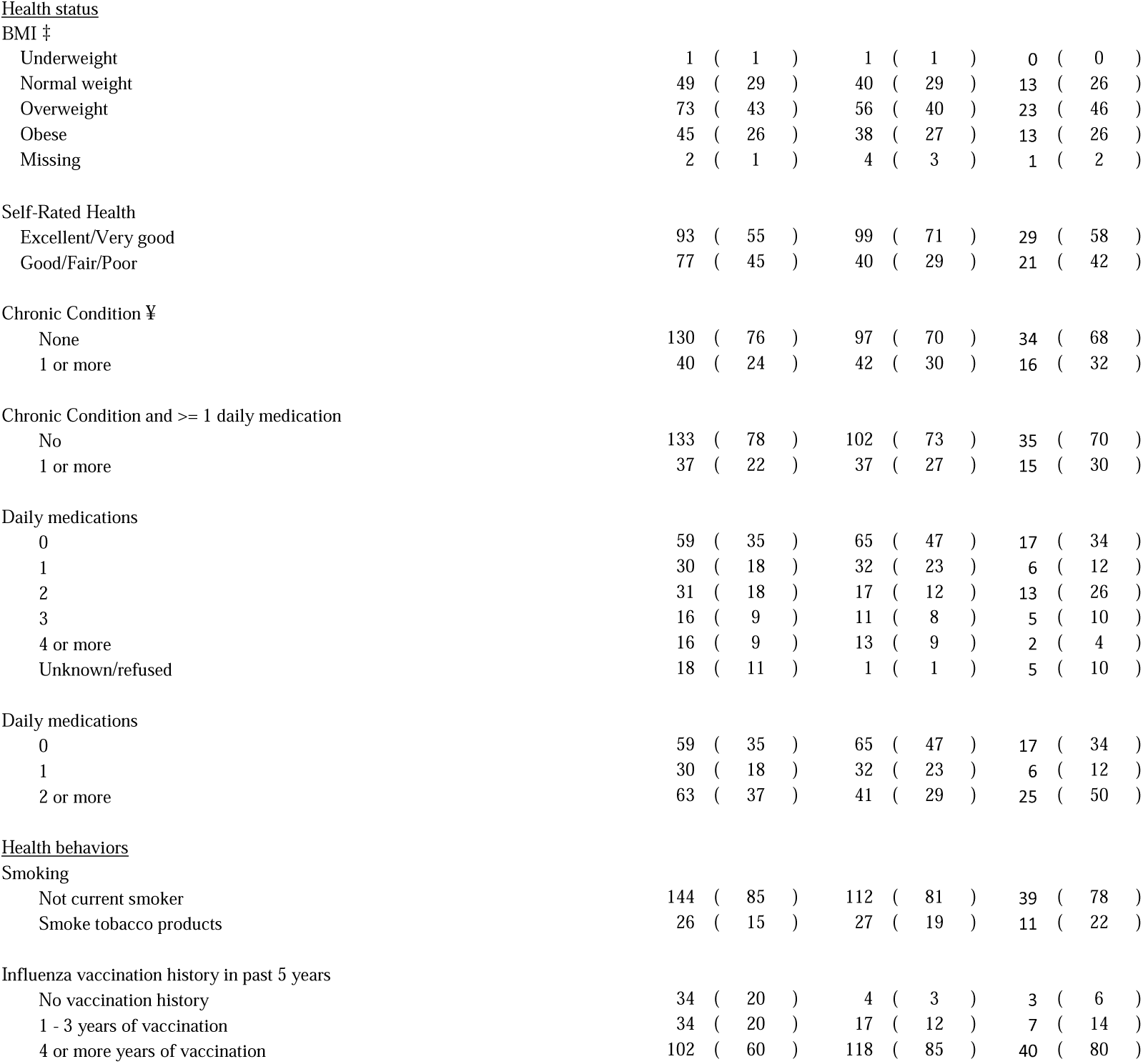
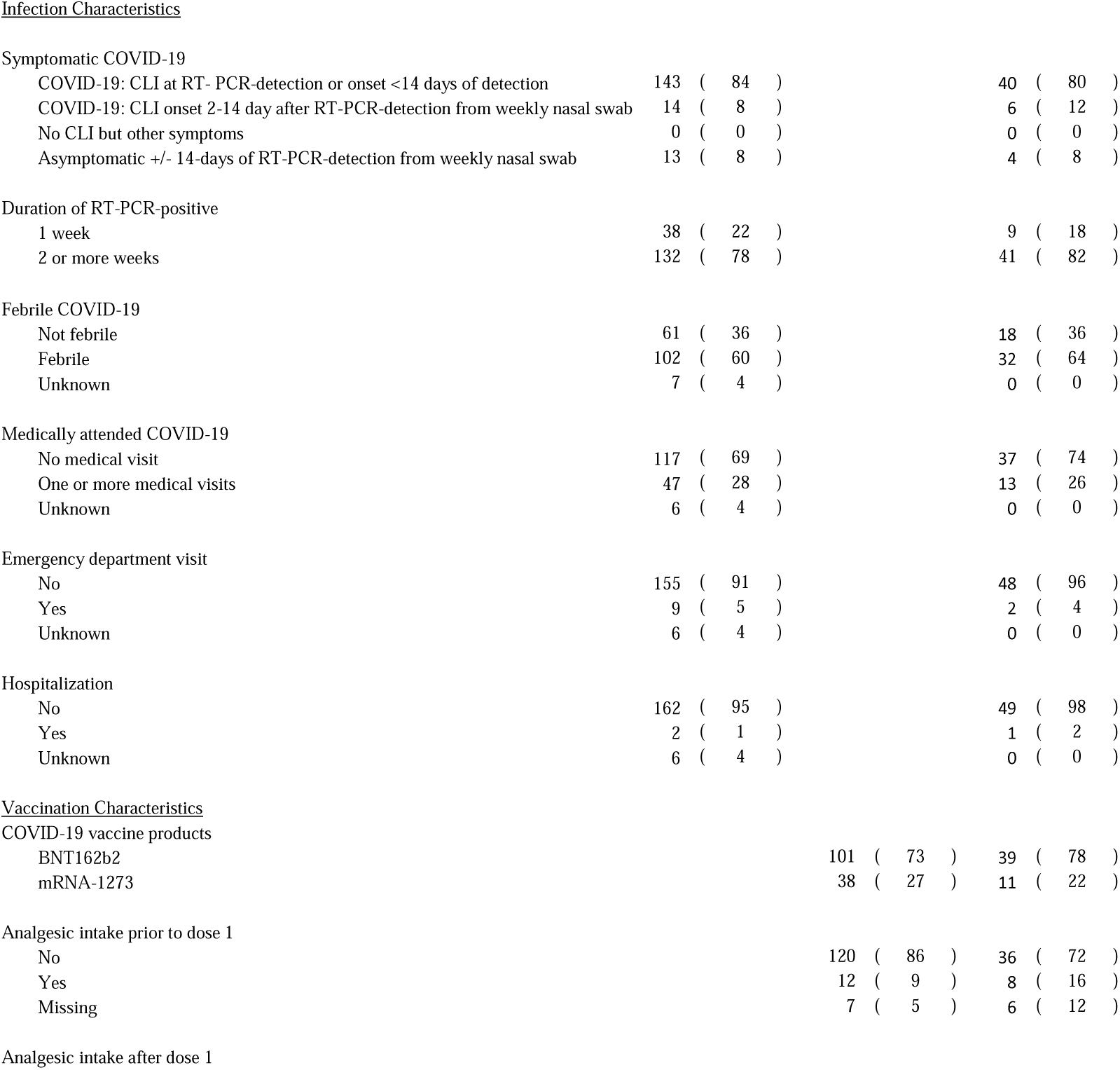
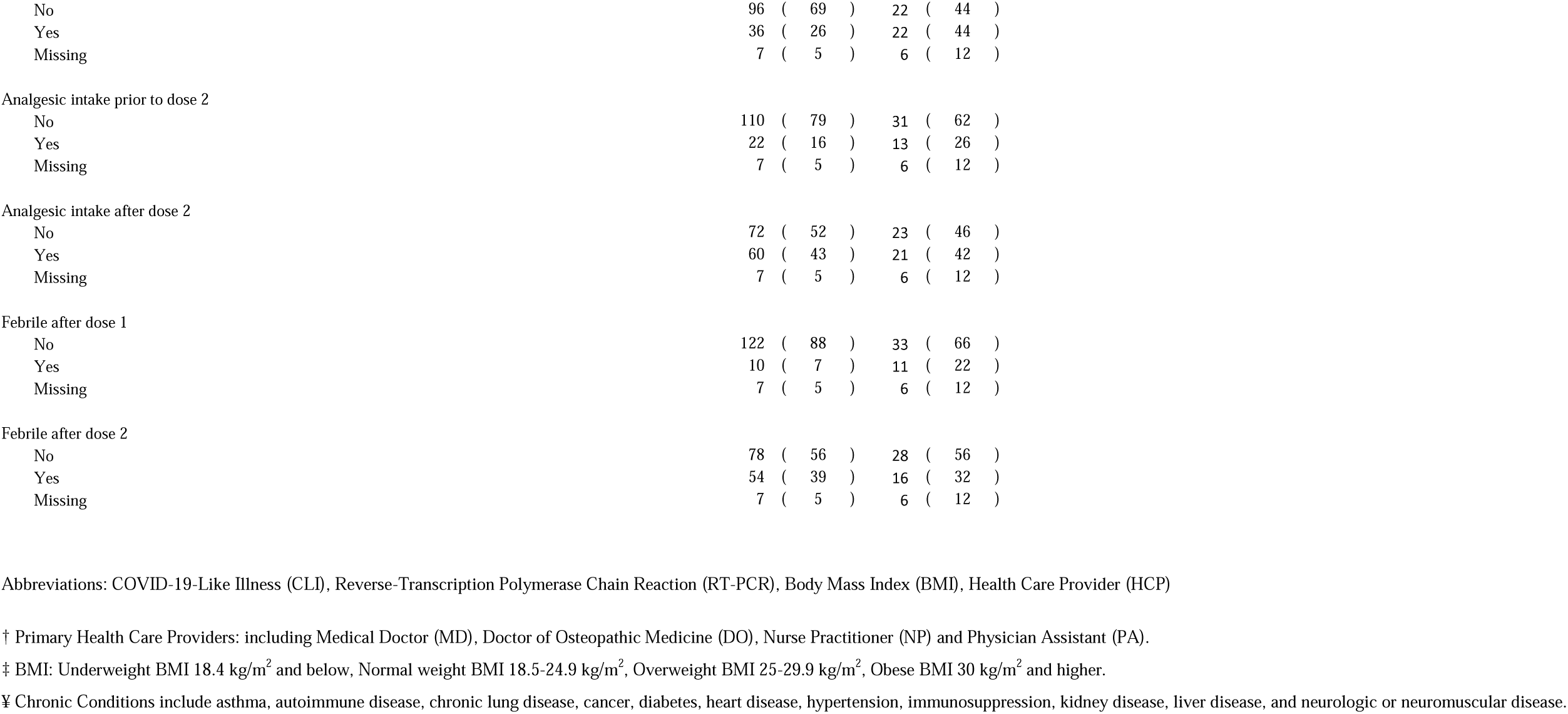
Participant socio-demographic and health characteristics, SARS-CoV-2 infection characteristics, and vaccination information.

### Response to SARS-CoV-2 infection

Of the 170 participants with RT-PCR-confirmed SARS-CoV-2 infection, most (92%) met symptom criteria for COVID-19; 8% were asymptomatic (Table 1). Most infections were associated with weekly positive swabs lasting for 2 or more weeks (78%) and fever (60%); 28% required medical care, 6% sought emergency care, and 1% required hospitalization. The median time from first positive swab collection to convalescent sera collection for infected participants was 30 days (IQR=26-37). Neutralizing antibodies were detected among 158 (93%) participants with evidence of infection (Table 2, Table S1). GMT post-infection, including those without detectable antibodies, was 1,003 (95%CI=766-1,315) (Table 2, Table S2). Neutralizing antibody GMT did not differ by SARS-CoV-2 infection characteristics, such as the presence or absence of febrile symptoms or duration of detectable virus (Table 2, Table S2); there was no association between nAb titer and highest viral RNA load isolated from nasal swabs collected during infection (p=0.78) (Figure S2). When compared to the 158 participants who developed nAb after SARS-CoV-2 infection, the 12 participants who did not develop nAb after SARS-CoV-2 infection were more likely to be smokers (5 of 12 compared to 21 of 158, respectively, (p = 0.02) (Table S1).

**Table 2.**
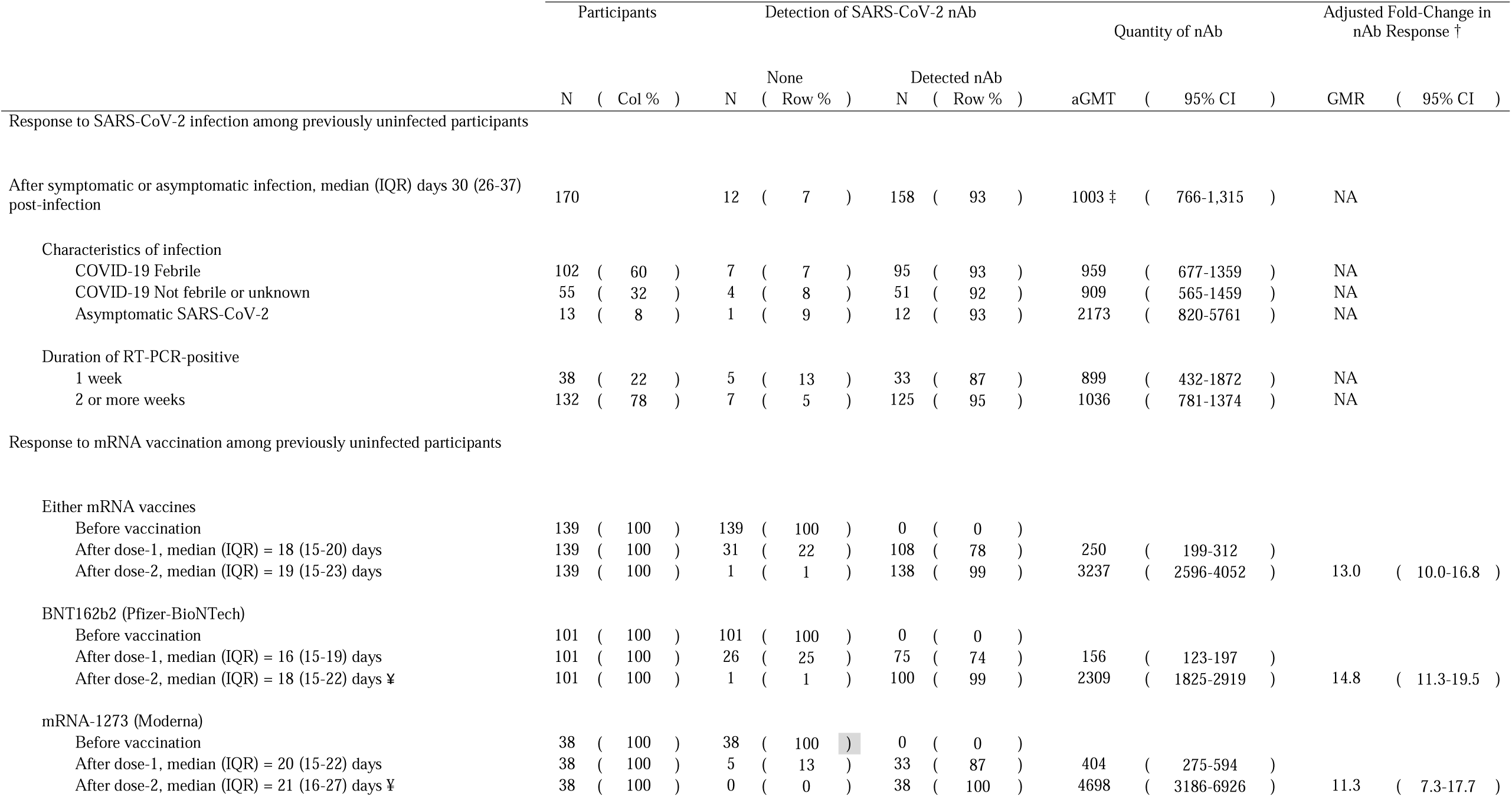
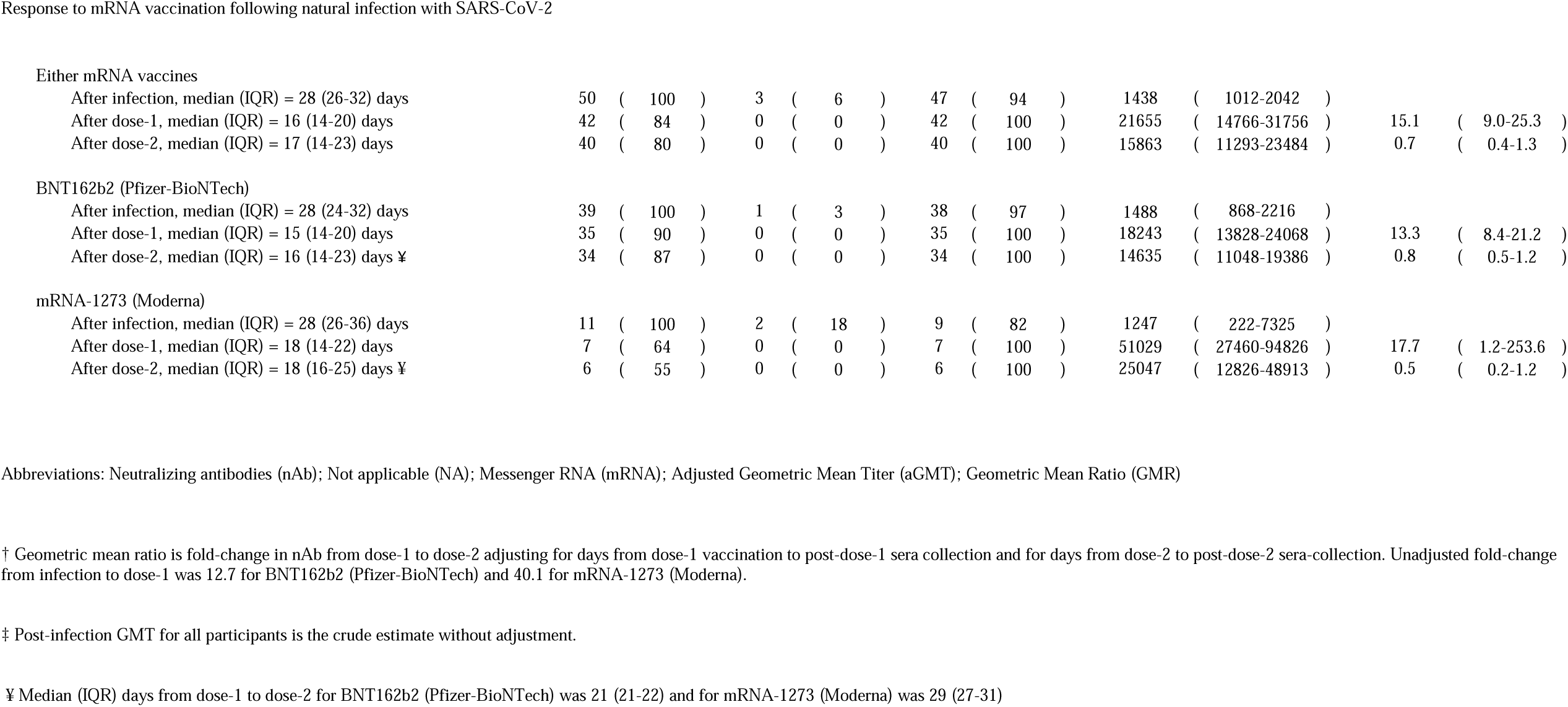
Detection and quantity of neutralizing antibodies (nAb) to SARS-CoV-2 following natural SARS-CoV-2 infection and vaccination with messenger RNA (mRNA) COVID-19 vaccines.

### Response to mRNA vaccination among previously uninfected participants

Sera were collected a median of 18 days (IQR=15-20 days) after mRNA vaccine dose-1 of either vaccine product (Table 2). Neutralizing antibodies were detected among 108 of 139 participants (78%) after dose-1 with a nAb GMT of 250 (95%CI=199-312) (Table 2). Neutralizing antibody GMT post-dose-1 was higher among those aged 18-49 years than those age 50 and older (p=0.05) and among those without chronic conditions (p=0.03) (Table S2).

Sera were collected a median of 19 days (IQR=15-23) after mRNA vaccine dose-2 of either vaccine product (Table 2). Neutralizing antibodies were detected among 138 of 139 participants (99%) with a nAb GMT of 3,237 (95%CI=2,596-4,052), a 13-fold (95%CI=10-16.8) increase in nAb titers (Table 2, Figure 2, Figure S1). Participants with a normal BMI (18.5-24.9 kg/m^2^) had lower nAb titers following dose-2 (p = 0.02) than those who were underweight, overweight, or obese (Table S2). Those with febrile symptoms following dose-2 had higher nAb GMT versus those who did not (3,587 versus 2,236, p=0.004), as did those who reported taking analgesics after dose 2 (3,312 versus 2,351, p = 0.03) (Table S2).

**Figure 2.**
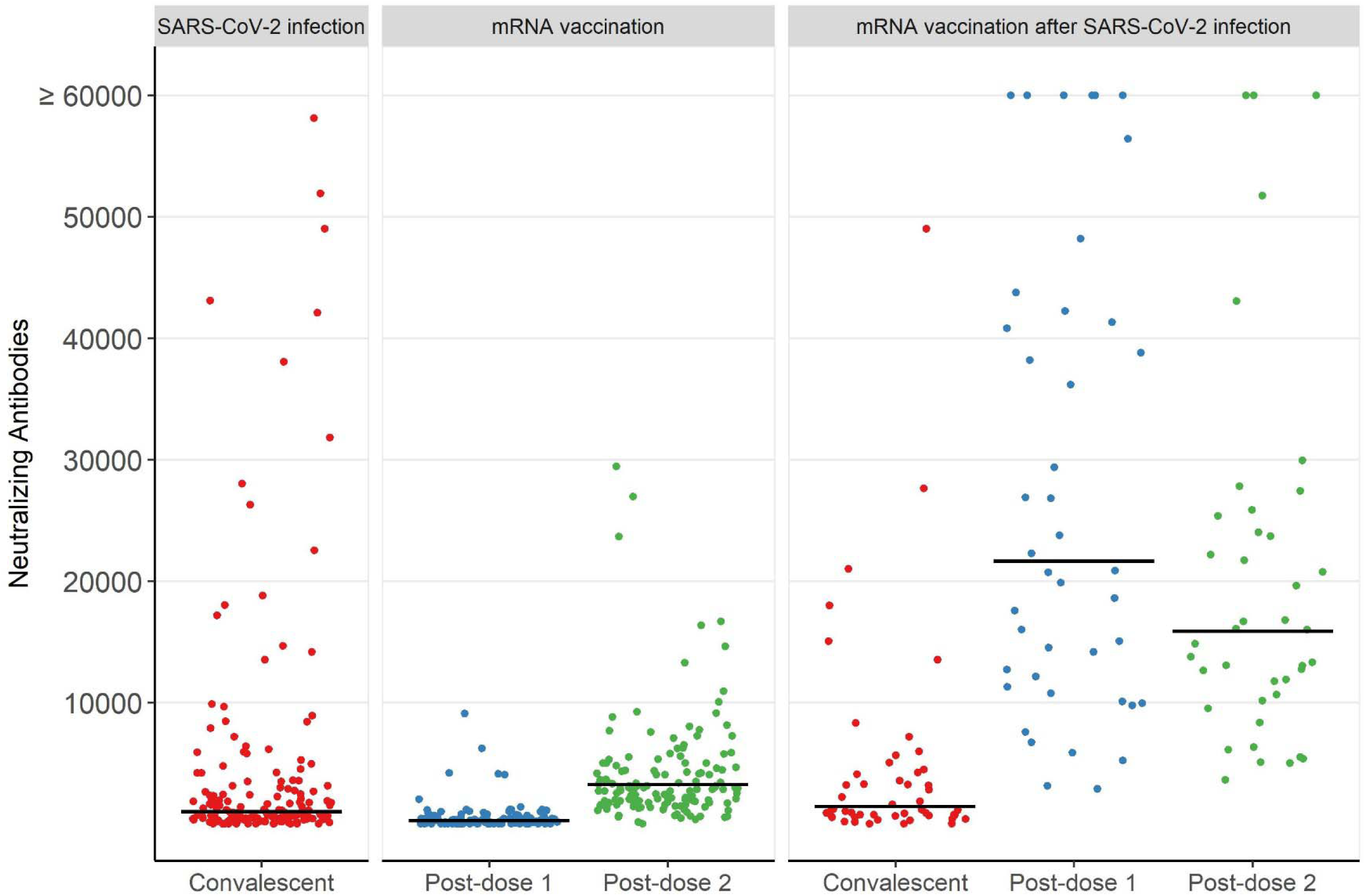
Neutralizing antibodies to pseudotype SARS-CoV-2 among sera convalescent to SARS-CoV-2 infection, approximately 14-21 days post-dose-1 and approximately 14-21 days post-dose-2 of messenger RNA COVID-19 vaccines among previously uninfected adults, and about 14-21 days post-doses-1 and 2 following vaccination among participants with prior positive SARS-CoV-2 test result. Legend: Dots are neutralizing antibody titers per participant. Line is geometric mean titer for that group.

Among the previously uninfected, response to the mRNA vaccines differed by product. Thirty-eight participants received mRNA-1273 vaccine and 101 participants received BNT162b2 vaccine (Table 2, Table S4). When comparing mRNA-1273 versus BNT162b2 recipients, mRNA-1273 recipients were more likely to be male (66% versus 44%) or a first responder (61% versus 4%) and less likely to be a primary health care provider (5% versus 37%) (Table S4) Among 38 recipients of the mRNA-1273 vaccine, nAb GMT after dose 1 was 404 (95% CI=275- 594) and increased 11-fold to 4,698 GMT (95%CI=3,186-6,926) following dose-2 (Table 2, Figure 3A). Among 101 recipients of the BNT162b2 vaccine, nAb GMT after dose-1 was 156 (95%CI=123-197) and increased 15-fold to 2,309 GMT (95% CI=1,825-2,919) following dose 2. Neutralizing antibody GMT was statistically higher for recipients of mRNA-1273 vaccine compared to recipients of BNT162b2 vaccine after dose-1 and after dose-2 (p <0.001) (Table 2, Figure 3A).

**Figure 3.**
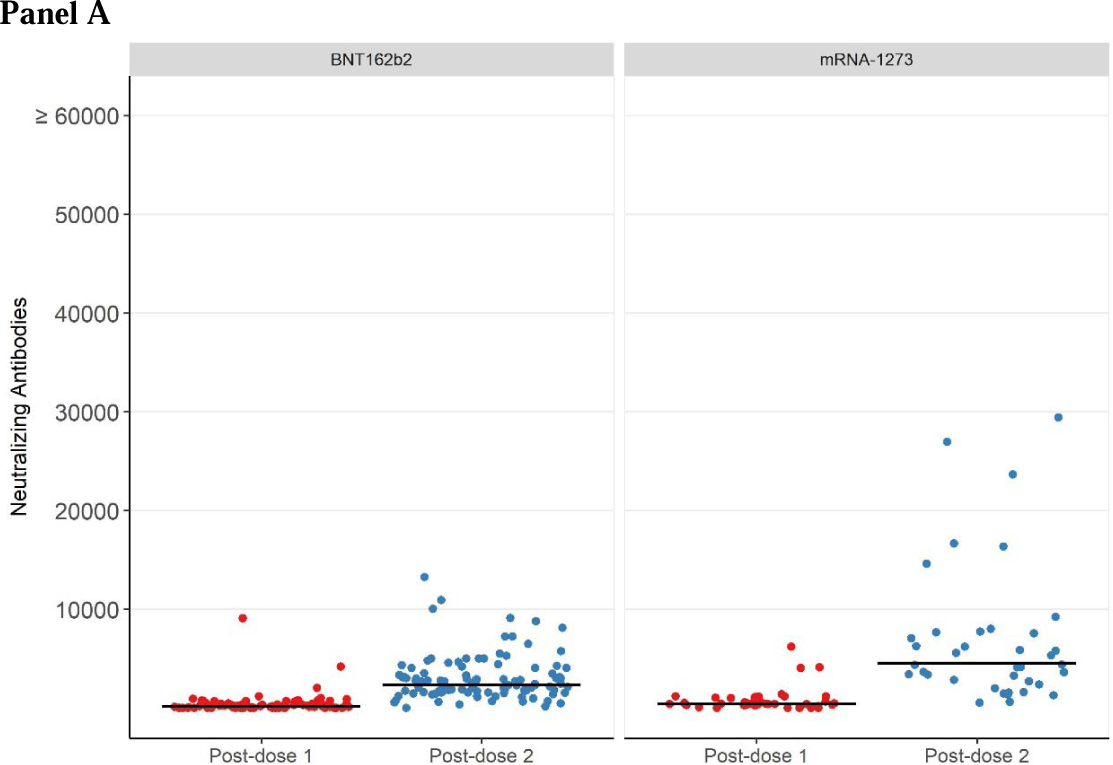
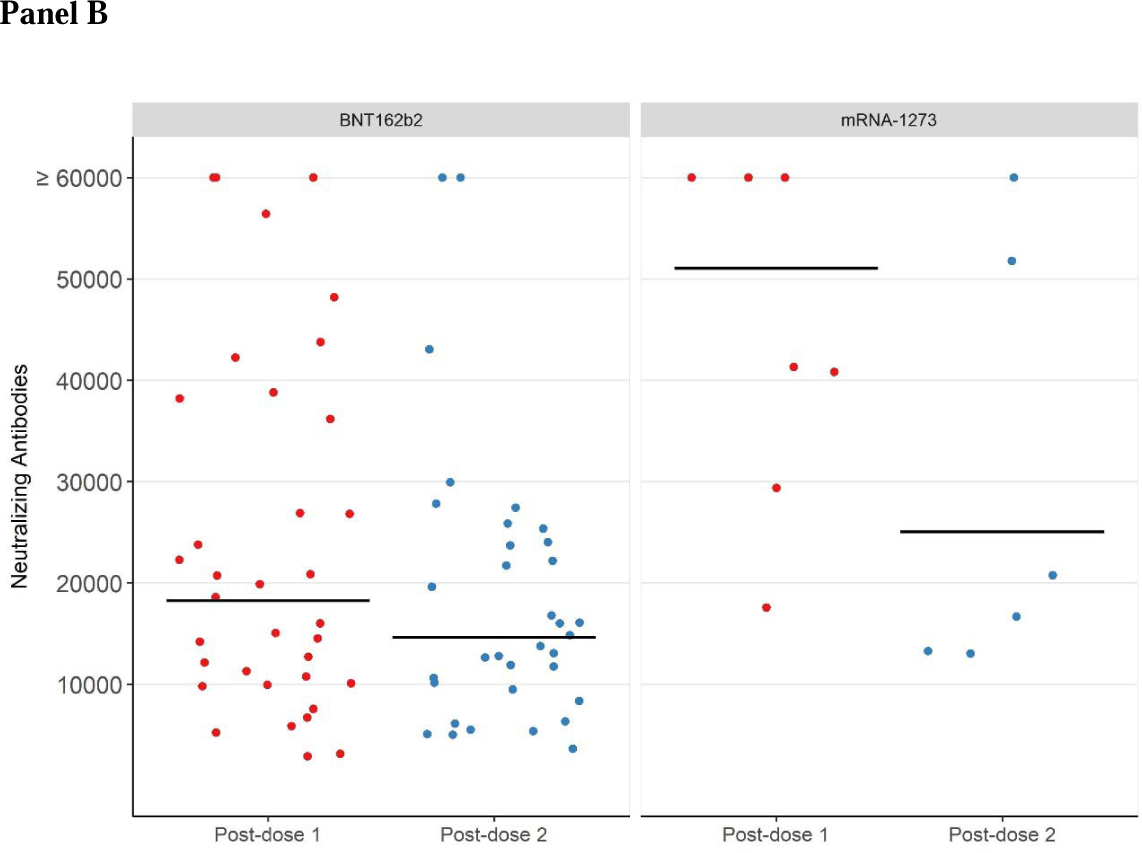
Neutralizing antibodies to pseudotype SARS-CoV-2 about 14-21 days post- dose 1 and 2 of BNT162b2 and mRNA-1273 COVID-19 vaccines among previously uninfected adults (A) and among participants with prior positive SARS-CoV-2 test result (B) Legend: Dots are neutralizing antibody titers per participant. Line is geometric mean titer (GMT) for that group.

### Response to mRNA vaccination following SARS-CoV-2 infection

Among the 50 participants who were infected with SARS-CoV-2, then subsequently vaccinated with either mRNA vaccine who had sera collected after dose-1 alone, dose-2 alone, or both doses (Figure S3), nAb GMT after infection and before vaccination was 1,438 (95% CI=1,012-2,042) (Table 2). Among the 42 participants with sera collected following mRNA vaccine dose-1, nAb increased 15.1-fold to nAb GMT of 21,655 (95% CI=14,766-31,756). Among the 40 participants with sera collected following mRNA vaccine dose-2, the nAb GMT was 15,863 (95% CI=11,293-23,484) which was statistically unchanged from the nAb GMT after vaccine dose-1. Participants with a prior positive SARS-CoV-2 test result with at least one chronic condition had significantly higher nAb response following vaccine dose-1 (p=0.02) and dose-2 (p=0.02) (Table S2). The pattern of nAb GMT increase was similar when analysis was restricted to the 32 participants with sera available for all 3 time points (Table S3). Neutralizing antibody GMT estimates following vaccine dose-1 were higher for recipients of the mRNA- 1,273 vaccine compared to the BNT162b2 vaccine, but confidence intervals were broad and overlapping (Table 2; Figure 3B).

### Comparisons across analytic samples

Taken together, the 139 previously uninfected participants who received mRNA vaccines achieved higher nAb titers (GMT =3,237, 95%CI=2,596-4,052) following a second vaccine dose compared to the nAb titers observed among 170 participants following SARS-CoV-2 infection (GMT=1,003, 95%CI=766-1,315) (Figure 2). The highest nAb GMT observed was among the 42 participants who were vaccinated after prior SARS-CoV-2 infection, achieving a nAb GMT of 21,655 (95%CI=14,766-31,756) following the first dose of an mRNA vaccine. The 3 participants who did not develop nAb after SARS-CoV-2 infection and had blood drawn after vaccination had a GMT rise comparable to the previously uninfected.

## DISCUSSION

In a prospective cohort of essential and frontline workers with no prior history of SARS-CoV-2 infection, the nAb activity to pseudotype SARS-CoV-2 measured 2-3 weeks following a second dose of mRNA COVID-19 vaccine was 13-fold higher than nAb activity measured following a single mRNA vaccine dose and over 3-fold higher than nAb activity measured 4-5 weeks after SARS-CoV-2 infection. Almost all study participants (99%) who received two doses of mRNA vaccine developed nAb, while no nAb were detected among 22% of participants following a single dose of an mRNA vaccine and 7% of adults following SARS-CoV-2 infection. The superior immunogenicity of two doses of mRNA vaccine compared to a single dose in real-world conditions is consistent with findings from efficacy trials [15, 16] and other recent observational studies [12]. However, our study is among the first to compare humoral immunogenicity to vaccination with humoral immune response to SARS-CoV-2 infection within the same cohort using common laboratory methods [17-19].

Vaccination had the greatest impact among adults with a prior positive SARS-CoV-2 test result; in this group of participants, nAb GMT was 21,655 following one dose of mRNA vaccine compared to a nAb GMT of 3,237 following two mRNA vaccine doses among previously uninfected adults. This is consistent with other findings that mRNA vaccine boosts humoral immune response to previous infection [20]. Like other studies, we found that a second dose did not further boost immune response among those with a previous infection [20-23]. Our results support the sufficiency of a single dose of mRNA vaccine to boost antibody response in most people with prior laboratory-confirmed infection. Moreover, the measurable increase in nAb GMT after at least one dose of vaccination after prior infection compared to the nAb GMT rise after infection alone supports the benefit of vaccination in the participants with a prior positive SARS-CoV-2 test result.

We found statistically significant differences in immunogenicity between mRNA vaccine products among participants with no prior infection. Neutralizing antibody GMT following a second dose of mRNA-1273 vaccine was 4,698compared to a nAb GMT of 2,309 following a second dose of BNT162b2 vaccine. Our study is among the first to compare the immunogenicity of the two mRNA vaccine products within the same cohort using the same laboratory methods [24]. Although finding a 2-fold difference between mRNA vaccine products in post-vaccination titers merits further investigation, it is also important to note that within this same cohort, vaccination with either mRNA vaccine product was associated with similar and high vaccine effectiveness against SARS-CoV-2 infection [25, 26]; similar product-specific vaccine efficacy estimates have been reported in other observational studies [27]. The GMT value necessary to provide protection has not been well described although titer correlates for protection have been proposed [28, 29].

Unlike other studies which have observed a correlation between more severe SARS-CoV-2 infection and higher antibody response [30-37], an association was not observed between febrile symptoms during infection, weeks of virus detection, or nasal viral RNA load and the development of nAb. Most prior studies focused on cases of severe COVID-19, often requiring hospitalization, while most of the infections examined in this study were mild and did not require hospitalization. Another reassuring finding was the lack of association between analgesic use before or after vaccination and reduced immunogenicity, which has been reported for some other vaccines [10]. In this study, analgesic use following a second mRNA vaccine dose was associated with higher nAb response. This may be related to our other finding that self-reported febrile symptoms following the second mRNA vaccine dose were also associated with higher antibody response. Similar associations between post-vaccination fever and immunogenicity have been noted for influenza vaccines [38].

Strengths of this multi-site study include routine weekly and illness testing irrespective of symptoms for SARS-CoV-2 infection by a molecular assay, collection of illness symptoms, documentation of viral load and duration of viral shedding, multimethod documentation of vaccination status, collection and processing of sera using common procedures, and the measurement of nAb to SARS-CoV-2 using a validated assay at a well-established and accredited clinical reference laboratory.

This study also has at least six limitations. First, nAb responses could only be examined within the time intervals of vaccination that occurred in our study. Inferences regarding antibody response following mRNA vaccine dose-1 are limited to the 2 to 3 weeks between vaccine doses, because most individuals had their second vaccine dose at the shortest recommended time interval. Second, serum-neutralizing antibodies were measured against USA-WA1/2020-spike pseudotype virus rather than with a live virus assay. However, previous studies have shown concordance between nonreplicating pseudotype neutralization and SARS-CoV-2 neutralization assays [39, 40]. Third, the nAb assays against USA-WA1/2020-spike pseudotype virus were performed by LabCorp using the PhenoSense® assay. The assay was validated before WHO standards were widely available, and the results are not reported in international units, therefore they are not directly translatable to assays performed by other laboratories. Fourth, cellular mediated immunogenicity could not be examined, which additionally contributes to clinical protection. In the same cohort, high vaccine effectiveness was noted in preventing SARS-CoV-2 infection starting 14 days after mRNA vaccine dose 1 [12]. Fifth, because of sparse data and limited sociodemographic and health heterogeneity, we were unable to fully examine or adjust for factors that may be associated with immune response to infection or vaccination. Data represented here was collected prior to the emergence of the SARS-CoV-2 lineage B.1.617.2 (Delta). Sixth, this study was conducted prior to the documentation of many infections after vaccination, however the measurement of nAb in previously infected and/or previously vaccinated individuals would provide information regarding the role of nAb in protective immunity and would be an important aim for future research.

In conclusion, in seronegative essential and frontline workers with no prior SARS-CoV-2 infection, two doses of mRNA vaccine resulted in higher nAb to SARS-CoV-2 than following one dose of vaccine or following SARS-CoV-2 infection alone, and a single dose of vaccine after SARS-CoV-2 infection resulted in higher nAb than two doses of mRNA COVID-19 vaccine in previously uninfected. Neutralizing antibody response differed by mRNA vaccine product and by history of prior infection, suggesting there are multiple dimensions on which the immune profile to SARS-CoV-2 varies in the general population.

## Data Availability

All data produced in the present work are contained in the manuscript

## All other authors

No conflicts.

## Acknowledgements

Danielle R. Hunt, Tyler C. Morrill, Brandon P. Poe, Brian Sokol, John Thacker, Tana Brummer, Andrea Bronaugh, James Carr, Hala Deeb, Sauma Doka, Claire Douglas, Kate Durocher, Tara Earl, Jini Etolue, Deanna Fleary, Isaiah Gerber, Kimberly Groover, Louise Hadden, Jenna Harder, Ed Hock, Keya Jacoby, Ryan Klein, Lindsay LeClair, Nancy McGarry, Peenaz Mistry, Steve Pickett, Khaila Prather, David Pulaski, Rajbansi Raorane, Alfredo Rodriguez-Nogues, Meghan Shea, Chao Zhou, Abt Associates; Michael E. Smith, Kempapura Murthy, Tnelda Zunie, Nicole Calhoun, Eric Hoffman, Martha Zayed, Joel Blais, Jason Ettlinger, Angela Kennedy, Natalie Settele, Rupande Patel, Elisa Priest, Jennifer Thomas, Madhava Beeram, Alejandro Arroliga, Baylor Scott & White Health; Julie Mayo Lamberte, Gregory Joseph, Josephine Mak, Monica Dickerson, Suxiang Tong, John Barnes, Eduardo Azziz-Baumgartner, Melissa L. Arvay, Preeta Kutty, Alicia M. Fry, Jill Ferdinands, Anthony Fiore, Aron Hall, Adam MacNeil, L. Clifford McDonald, Mary Reynolds, Sue Reynolds, Stephanie Schrag, Nong Shang, Robert Slaughter, Matthew J. Stuckey, Jennifer Verani, Vic Veguilla, Rose Wang, Bao-Ping Zhu, William Brannen, Stephanie Bialek, CDC; Yolanda Prado, Daniel Sapp, Mi Lee, Chris Eddy, Matt Hornbrook, Donna Eubanks, Danielle Millay, Dorothy Kurdyla, Kristin Bialobok, Ambrosia Bass, Kristi Bays, Kimberly Berame, Cathleen Bourdoin, Carlea Buslach, Jennifer Gluth, Kenni Graham, Tarika Holness, Enedina Luis, Abreeanah Magdaleno, DeShaun Martin, Joyce Smith-McGee, Martha Perley, Sam Peterson, Aaron Piepert, Krystil Phillips, Joanna Price, Sperry Robinson, Katrina Schell, Emily Schield, Natosha Shirley, Anna Shivinsky, Britta Torgrimson-Ojerio, Brooke Wainwright, Shawn Westaway, Kaiser Permanente Northwest; Angela Hunt, Jessica Lundgren, Karley Respet, Jennifer Viergutz, Daniel Stafki, St. Luke’s Regional Health Care System; Zoe Baccam, Drew Baldwin, Genesis Barron, Dimaye Calvo, Esteban Cardona, Adam Carl, Andrea Carmona, Alissa Coleman, Emily Cooksey, David Dawley, Stacy Delgado, Kiara Earley, Katherine Ellingson, Natalie Giroux, Anna Giudici, Sofia Grijalva, Allan Guidos, Hanna Hanson, Adrianna Hernandez, James Hollister, Theresa Hopkins, Rezwana Islam, Gabriella Jimenez, Krystal Jovel, Olivia Kavanagh, Karla Ledezma, Sally Littau, Amelia Lobos, James Lopez, Veronica Lugo, Jeremy Makar, Taylor Maldonado, Enrique Marquez, Christina Mortensen, Allyson Munoz, Flavia Nakayima Miiro, Sandra Norman, Assumpta Nsengiyunva, Kennedy Obrien, Jonathan Perez Leyva, Celia Pikowski, Cynthia Porter, Patrick Rivers, James Romine, Alexa Roy, Jennifer Scott, Saskia Smidt, Gianna Taylor, Isabella Terrazas, Tahlia Thompson, Heena Timisina, Jennifer Uhrlaub, Erica Vanover, Mandie White, and April Yingst, University of Arizona; Carlos Silvera, Cynthia Beaver, Roger Noriega, Alexandrea Cruz, Damena Gallimore-Wilson, Rachel Reyes, Christian Rojas, Catalina Gonzalez, Simi Oduwole, -Umana, Addison Testoff, Alex Stewart, Kemi Ogunsina, Aimee Green, Johanna Garibaldi, Nathaly Suarez, Olga Carrera, Hannah KlingUniversity of Miami; Matthew Bruner, Rachel Brown, Jenna Praggastis, Marcus Stucki, Arlyne Arteaga, Riley Campbell, Madeleine Smith, Adriele Fugal, Maya Wheeler, Gretchen Maughan, Allana Soriano, Nikki Gallacher, Anika Dsouza, Lauren Anderson, Jenna Vo, Trevor Stubbs, Iman Ibrahim, Tristen Forbes, Taryn Hunt-Smith, Ryder Jordin, Michael Langston, Timina Powaukee, Daniel Dawson, Kathy Tran, Fiona Tsang, Hannah Whiting, Emilee Eden, Braydon Black, Christina Pick, Madison Tallman, Chapman Cox, Derrick Wong, Camie Schaefer, Andrew L. Phillips, Kurt T. Hegmann, Jeanmarie Mayer, Joseph Stanford, University of Utah; Marilyn J. Odean, Whiteside Institute for Clinical Research; Allen Bateman, Erik Reisdorf, Kyley Guenther, Erika Hanson, Wisconsin State Laboratory of Hygiene; Labcorp, Monogram Biosciences.

