## Supplement Table 1 for "Neutralizing Antibody Response to Pseudotype SARS-CoV-2 Differs between mRNA-1273 and BNT162b2 COVID-19 Vaccines and by History of SARS-CoV-2 Infection"

SUPPLEMENTARY APPENDIX

METHODS

Laboratory Methods:

**Neutralizing antibody assay:**

A SARS-CoV-2 pseudovirus was produced that expresses spike protein of the Wuhan reference virus USA-WA1/2020. Methods used to create this pseudovirus can be found in Peluso et al., 2021, Cell Reports 36, 109518 [14]. Sera specimens were heat inactivated and diluted 1:40 in cell culture medium. Neutralizing antibody titers were determined by generating serial 10-fold dilutions of participant samples that were incubated with the SARS-CoV-2 pseudovirus. The results of the SARS-CoV-2 LabCorp PhenoSense ^®^ neutralizing antibody assay, an in-house laboratory developed test manufactured by Monogram Biosciences in South San Francisco, CA, [14], are reported as a ID50 titer and as positive/negative based on a pre-defined (>50% inhibition at 1:40 dilution) dilution cutoff. To confirm that the neutralizing antibody is specific to SARS-CoV-2, each sera sample is tested using a specificity control, in this case Avian influenza pseudovirus H10N7, which is run in parallel with the SARS-CoV-2 pseudovirus. The assay is performed by generating a serial dilution of the serum. The dilution is stamped to two assay plates. The SARS-CoV-2 pseudovirus is added to the first assay plate, the specificity control Avian influenza pseudovirus H10N7 is added to the second assay plate. The SARS-CoV-2 nAb activity observed in the first plate is only valid when the specificity control (H10N7) activity is negative. Positive and negative control plasma for SARS-CoV-2 neutralization antibodies are included on each assay plate along with participant sera samples.

**Study Population:**

The time period considered was from the time of enrollment through March 15th, 2021, when sera were selected to be sent for analysis. All participants were required to be SARS-CoV-2 naïve at enrollment by self-report and Ortho VITROS Anti-SARS-CoV-2 Total IgG Assay (Ortho Clinical Diagnostics) ELISA. Individuals who tested positive for SARS-CoV-2 by RT-PCR nasal swab during the study were asked to provide a convalescent serum specimen 28 days after their first positive test result. All participants who were vaccinated against COVID-19 were asked to submit a serum sample 14 days after each dose of vaccine. Participants receiving the Johnson and Johnson vaccine were excluded in analysis due to the small number of recipients. Participants with no prior reported infection but with a positive pre-infection or pre-vaccination nAb result were removed from analysis. Of the 3,975 participants in the prospective cohort study, a total of 497 participants were identified for the analytic populations from August 15, 2020 through March 15, 2021.

Response to SARS-CoV-2 infection

To be included in the infection-only analysis population, participants needed to have a blood draw demonstrating negative nAb status at enrollment, a positive swab for SARS-CoV-2 during the study, as well as a convalescent blood draw approximately 28 days after the first positive, prior to any SARS-CoV-2 vaccination. Of the 337 participants who has a positive swab for SARS-CoV-2 during the study, 170 were confirmed to have a negative antibody test prior to infection and had serum drawn approximately 28 days after infection, before vaccination.

Response to mRNA vaccination among previously uninfected participants

To be included in the analysis population for previously uninfected, post-vaccination, participants needed to have a blood draw demonstrating negative nAb status at enrollment and all tested blood draw prior to vaccination. They could not have had a positive swab for SARS-CoV-2 and could not have any indication of SARS-CoV-2 infection prior to receiving both doses of mRNA vaccine. Participants also had to have submitted serum samples approximately 14 days after both doses of vaccine within the study period. Of the 327 participants meeting criteria, a random sample of 140 was selected for nAb analysis, stratified by age and sex. The number 140 was chosen because of a limited number of available tests.

Response to mRNA vaccination following infection with SARS-CoV-2

Participants in this analysis population had to meet all the criteria for the analysis population noted above as the “Response to SARS-CoV-2 infection”. Additionally, they needed to subsequently receive both doses of mRNA vaccine and have serum drawn approximately 14 days after at least one of the doses. Of the 170 in the “Response to SARS-CoV-2 infection” analysis population, 50 participants were vaccinated within the study period and had blood drawn after at least one of the doses. 10 participants had blood drawn only after dose 1, 8 participants after only dose 2, and 32 participants had blood drawn after both doses 1 and 2.

**Statistical Methods:**

Statistical analysis

### Response to SARS-CoV-2 infection among previously uninfected participants:

Overall geometric mean titer (GMT) and confidence intervals (CI) were calculated by a one-sample t-test. GMTs and CIs by infection characteristics were calculated with two-sample t-tests for dichotomous variables, one-way analysis of variance for multicategory variables, and analysis of covariance for continuous variables. We considered adjusted models including days from first SARS-CoV-2 RT-PCR positive sample to serum blood draw and as well as socio demographic and health characteristics. No potential covariates met the criteria of adjusting estimates by at least 5%.

### Response to mRNA vaccination among previously uninfected participants:

GMTs and CIs were estimated with a mixed effects model with a repeated measure for nAb following dose 1 and dose 2, main effects of blood draw (post dose 1 or 2) and vaccine product and their interaction. No potential covariates met the criteria of adjusting estimates by at least 5%. GMTs are back transformed least squares means and geometric mean fold rises are back transformed difference of means.

Secondary models additionally included fever after vaccination, analgesics before vaccination, and analgesics after vaccination to estimate their impact on GMT.

### Response to mRNA vaccination following infection with SARS-CoV-2:

GMTs and CIs after infection was calculated with a one sample t-test. GMTs and CIs with the same model as the vaccination among previously uninfected group. We considered using log_10_ nAb titers after infection as a covariate but it was insignificant and did not meet the >5% criteria. No additional covariates were tested due to lack of power with the small sample size.

Not all subjects had both a dose 1 and a dose 2 blood draw. As a sensitivity analysis we reran the model on the subset of subjects who had both draws.

Figure S1. Neutralizing antibodies to pseudotype SARS-CoV-2 on a log^10^ scale (Y-axis) among sera convalescent to SARS-CoV-2infection (panel 1), mRNA vaccination among previously uninfected adults (panel 2), and mRNA vaccination after SARS-CoV-2l infection (panel 3).

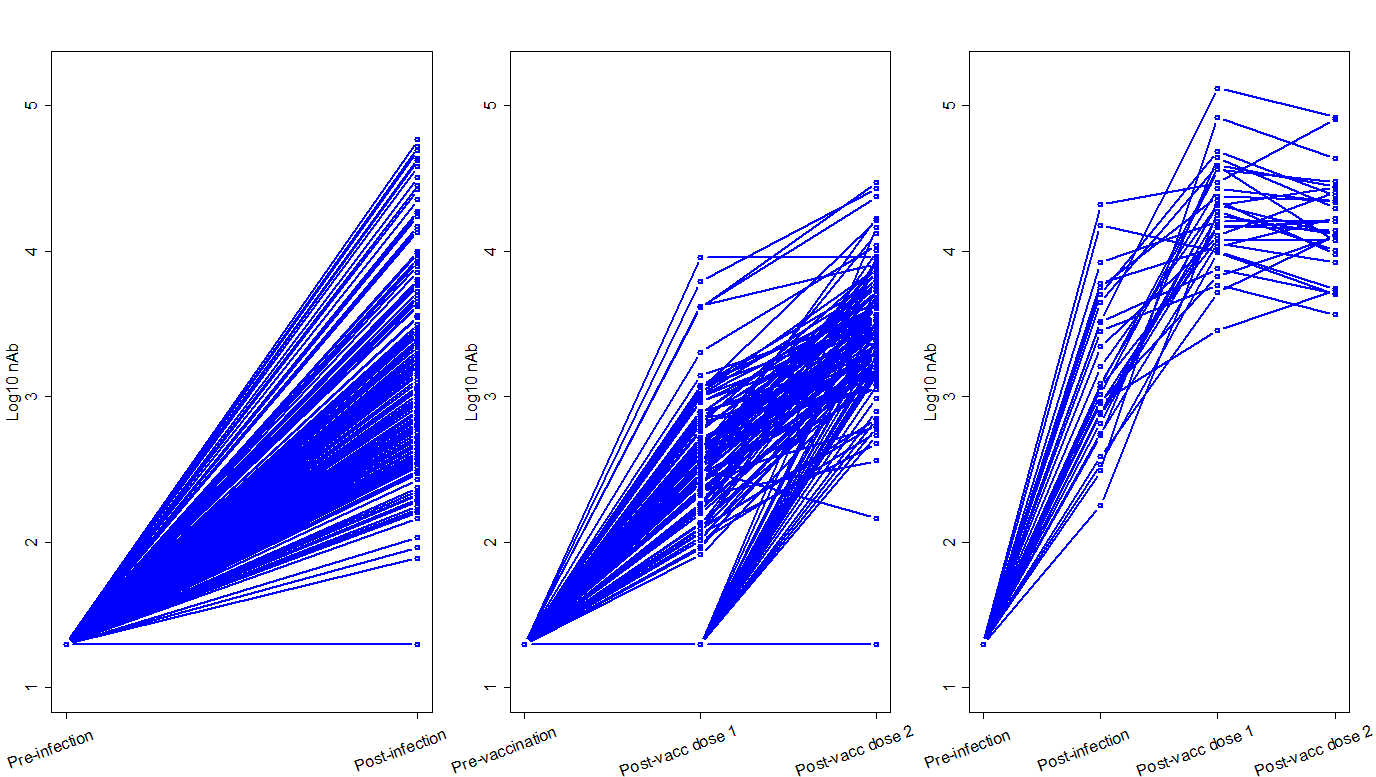

Figure S2. Neutralizing antibodies to pseudotype SARS-CoV-2 among sera convalescent to SARS-CoV-2 infection (Y-axis) by viral RNA load from nasal specimens collected at infection

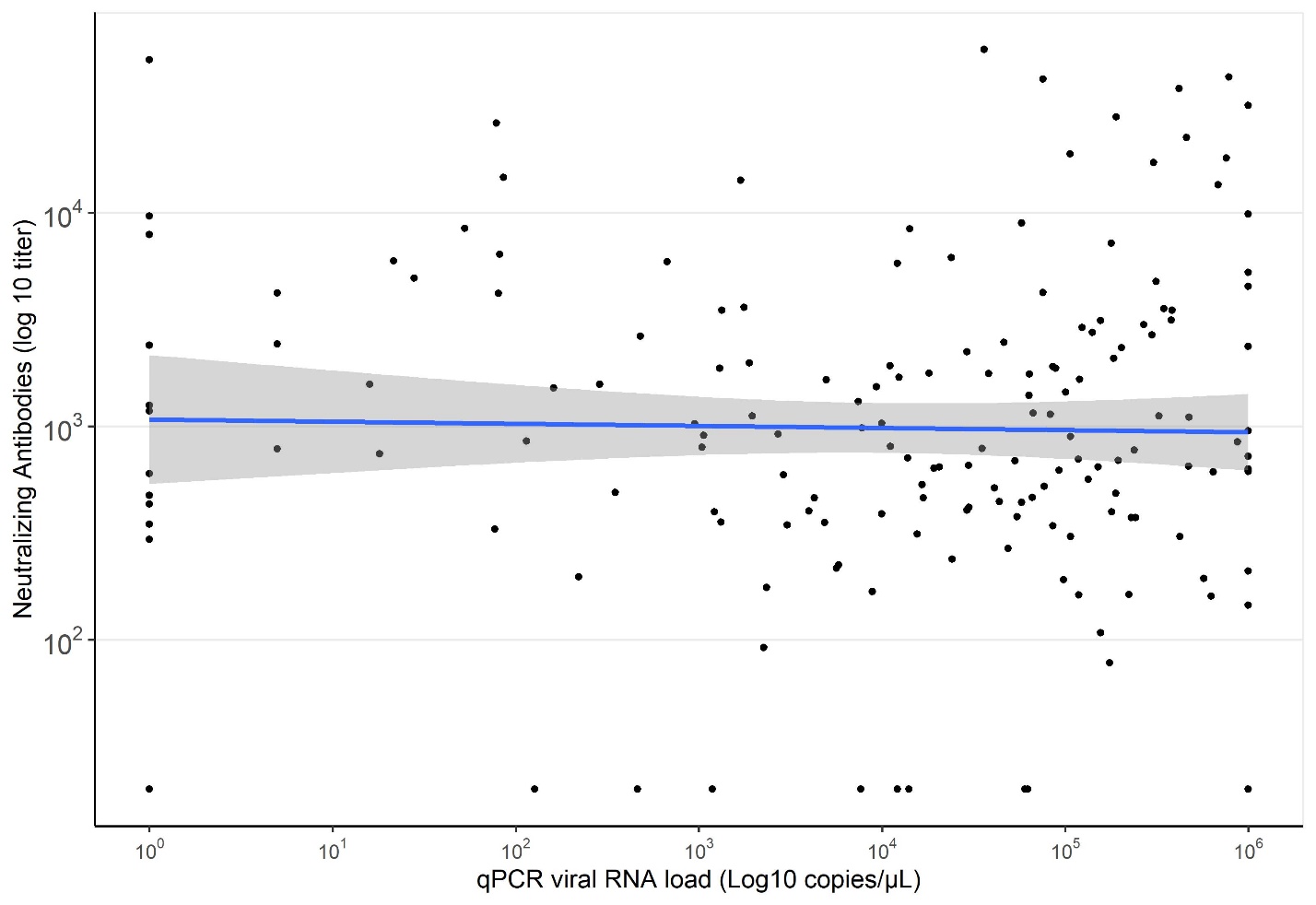

Figure S3. Serum Draws by Vaccine Dose. Participants previously infected with SARS CoV-2 who had blood drawn after one or both doses of mRNA SARS CoV-2 vaccine.

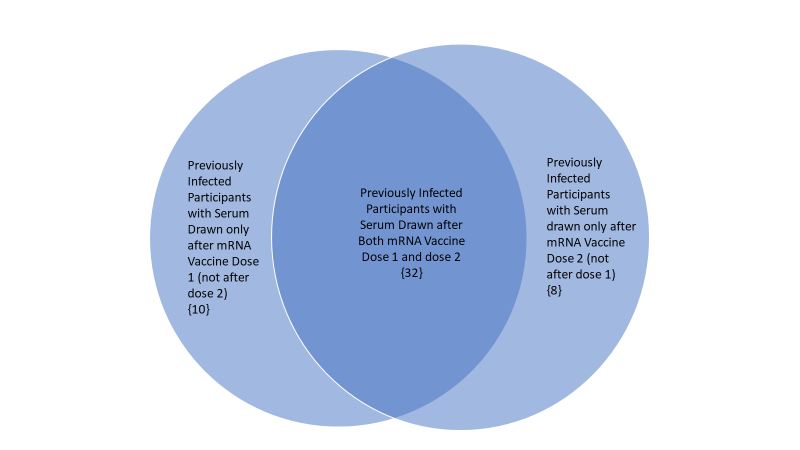

| **Supplementary Table 1. Characteristics of participants who did and did not have detectable neutralizing antibodies (nAb) to SARS-CoV-2 following natural SARS-CoV-2 infection and vaccination with the first dose of messenger RNA (mRNA) COVID-19 vaccines** | | | | | | | | | | | | | | | | | | | | | | | |
| --- | --- | --- | --- | --- | --- | --- | --- | --- | --- | --- | --- | --- | --- | --- | --- | --- | --- | --- | --- | --- | --- | --- | --- |
|  | **Response to RT-PCR confirmed infection; N=170** | | | | | | | | | | |  | **Response to vaccine dose 1; N=139** | | | | | | | | | | |
|  | **nAb positive after infection** | | | |  | **nAb negative after infection** | | | |  |  | **nAb positive after vaccine dose one** | | | | |  | **nAb negative after vaccine dose one** | | | |  |  |
|  | N | ( | Col. % | ) |  | N | ( | Col. % | ) |  | p-value |  | N | ( | Col. % | ) |  | N | ( | Col. % | ) |  | p-value |
| All participants (row %) | 158 | ( | 93 | ) |  | 12 | ( | 7 | ) |  |  |  | 108 | ( | 78 | ) |  | 31 | ( | 22 | ) |  |  |
| Socio-demographic characteristics |  |  |  |  |  |  |  |  |  |  |  |  |  |  |  |  |  |  |  |  |  |  |  |
| Cohort location |  |  |  |  |  |  |  |  |  |  | 0.298 |  |  |  |  |  |  |  |  |  |  |  | 0.104 |
| Phoenix, AZ | 19 | ( | 12 | ) |  | 1 | ( | 8 | ) |  |  |  | 0 | ( | 0 | ) |  | 0 | ( | 0 | ) |  |  |
| Tucson, AZ | 42 | ( | 27 | ) |  | 4 | ( | 33 | ) |  |  |  | 3 | ( | 3 | ) |  | 0 | ( | 0 | ) |  |  |
| Other, AZ | 3 | ( | 2 | ) |  | 0 | ( | 0 | ) |  |  |  | 0 | ( | 0 | ) |  | 0 | ( | 0 | ) |  |  |
| Miami, FL | 13 | ( | 8 | ) |  | 4 | ( | 33 | ) |  |  |  | 7 | ( | 6 | ) |  | 0 | ( | 0 | ) |  |  |
| Duluth, MN | 33 | ( | 21 | ) |  | 1 | ( | 8 | ) |  |  |  | 60 | ( | 56 | ) |  | 25 | ( | 81 | ) |  |  |
| Portland, OR | 8 | ( | 5 | ) |  | 0 | ( | 0 | ) |  |  |  | 35 | ( | 32 | ) |  | 5 | ( | 16 | ) |  |  |
| Temple, TX | 24 | ( | 15 | ) |  | 2 | ( | 17 | ) |  |  |  | 1 | ( | 1 | ) |  | 1 | ( | 3 | ) |  |  |
| Salt Lake City, UT | 15 | ( | 9 | ) |  | 0 | ( | 0 | ) |  |  |  | 2 | ( | 2 | ) |  | 0 | ( | 0 | ) |  |  |
| Sex |  |  |  |  |  |  |  |  |  |  | 0.774 |  |  |  |  |  |  |  |  |  |  |  | 0.572 |
| Female | 71 | ( | 45 | ) |  | 6 | ( | 50 | ) |  |  |  | 53 | ( | 49 | ) |  | 17 | ( | 55 | ) |  |  |
| Male | 86 | ( | 54 | ) |  | 6 | ( | 50 | ) |  |  |  | 55 | ( | 51 | ) |  | 14 | ( | 45 | ) |  |  |
| Age (Years) |  |  |  |  |  |  |  |  |  |  | 0.339 |  |  |  |  |  |  |  |  |  |  |  | 0.084 |
| 18-49 | 113 | ( | 72 | ) |  | 7 | ( | 58 | ) |  |  |  | 77 | ( | 71 | ) |  | 17 | ( | 55 | ) |  |  |
| ≥50 | 45 | ( | 28 | ) |  | 5 | ( | 42 | ) |  |  |  | 31 | ( | 29 | ) |  | 14 | ( | 45 | ) |  |  |
| Race |  |  |  |  |  |  |  |  |  |  | 1.000 |  |  |  |  |  |  |  |  |  |  |  | 0.300 |
| White | 146 | ( | 92 | ) |  | 11 | ( | 92 | ) |  |  |  | 97 | ( | 90 | ) |  | 30 | ( | 97 | ) |  |  |
| Other | 12 | ( | 8 | ) |  | 1 | ( | 8 | ) |  |  |  | 11 | ( | 10 | ) |  | 1 | ( | 3 | ) |  |  |
| Ethnicity |  |  |  |  |  |  |  |  |  |  | 0.693 |  |  |  |  |  |  |  |  |  |  |  | 0.575 |
| Hispanic/Latinx | 27 | ( | 17 | ) |  | 1 | ( | 8 | ) |  |  |  | 4 | ( | 4 | ) |  | 0 | ( | 0 | ) |  |  |
| Other | 131 | ( | 83 | ) |  | 11 | ( | 92 | ) |  |  |  | 104 | ( | 96 | ) |  | 31 | ( | 100 | ) |  |  |
| Occupation |  |  |  |  |  |  |  |  |  |  | 0.228 |  |  |  |  |  |  |  |  |  |  |  | 0.384 |
| Primary HCP | 20 | ( | 13 | ) |  | 1 | ( | 8 | ) |  |  |  | 28 | ( | 26 | ) |  | 11 | ( | 35 | ) |  |  |
| Nurses and other allied HCP | 64 | ( | 41 | ) |  | 2 | ( | 17 | ) |  |  |  | 55 | ( | 51 | ) |  | 17 | ( | 55 | ) |  |  |
| First Responders | 60 | ( | 38 | ) |  | 7 | ( | 58 | ) |  |  |  | 24 | ( | 22 | ) |  | 3 | ( | 10 | ) |  |  |
| Essential and other frontline | 14 | ( | 9 | ) |  | 2 | ( | 17 | ) |  |  |  | 1 | ( | 1 | ) |  | 0 | ( | 0 | ) |  |  |
| Health status |  |  |  |  |  |  |  |  |  |  |  |  |  |  |  |  |  |  |  |  |  |  |  |
| BMI |  |  |  |  |  |  |  |  |  |  | 0.936 |  |  |  |  |  |  |  |  |  |  |  | 0.302 |
| Underweight | 1 | ( | 1 | ) |  | 0 | ( | 0 | ) |  |  |  | 1 | ( | 0 | ) |  | 0 | ( | 0 | ) |  |  |
| Normal weight | 46 | ( | 29 | ) |  | 3 | ( | 25 | ) |  |  |  | 30 | ( | 1 | ) |  | 1 | ( | 3 | ) |  |  |
| Overweight | 67 | ( | 42 | ) |  | 6 | ( | 50 | ) |  |  |  | 45 | ( | 28 | ) |  | 9 | ( | 29 | ) |  |  |
| Obese | 42 | ( | 27 | ) |  | 3 | ( | 25 | ) |  |  |  | 26 | ( | 42 | ) |  | 12 | ( | 39 | ) |  |  |
| Missing | 2 | ( | 1 | ) |  | 0 | ( | 0 | ) |  |  |  | 6 | ( | 24 | ) |  | 0 | ( | 0 | ) |  |  |
| Self-Rated Health |  |  |  |  |  |  |  |  |  |  | 0.381 |  |  |  |  |  |  |  |  |  |  |  | 1 |
| Excellent/Very good | 88 | ( | 56 | ) |  | 5 | ( | 42 | ) |  |  |  | 2 | ( | 2 | ) |  | 0 | ( | 0 | ) |  |  |
| Good/Fair/Poor | 70 | ( | 44 | ) |  | 7 | ( | 58 | ) |  |  |  | 104 | ( | 96 | ) |  | 31 | ( | 100 | ) |  |  |
| Chronic Condition |  |  |  |  |  |  |  |  |  |  | 1.000 |  |  |  |  |  |  |  |  |  |  |  | 0.042 |
| None | 121 | ( | 77 | ) |  | 9 | ( | 75 | ) |  |  |  | 79 | ( | 73 | ) |  | 16 | ( | 52 | ) |  |  |
| 1 or more | 37 | ( | 23 | ) |  | 3 | ( | 25 | ) |  |  |  | 27 | ( | 25 | ) |  | 15 | ( | 48 | ) |  |  |
| Chronic Condition and >= 1 daily medication |  |  |  |  |  |  |  |  |  |  | 0.725 |  |  |  |  |  |  |  |  |  |  |  | 0.200 |
| None | 124 | ( | 78 | ) |  | 9 | ( | 75 | ) |  |  |  | 81 | ( | 75 | ) |  | 19 | ( | 61 | ) |  |  |
| 1 or more | 34 | ( | 22 | ) |  | 3 | ( | 25 | ) |  |  |  | 25 | ( | 23 | ) |  | 12 | ( | 39 | ) |  |  |
| Daily medications |  |  |  |  |  |  |  |  |  |  | 0.762 |  |  |  |  |  |  |  |  |  |  |  | 0.494 |
| 0 | 56 | ( | 11 | ) |  | 3 | ( | 25 | ) |  |  |  | 51 | ( | 47 | ) |  | 12 | ( | 39 | ) |  |  |
| 1 | 27 | ( | 35 | ) |  | 3 | ( | 25 | ) |  |  |  | 23 | ( | 21 | ) |  | 9 | ( | 29 | ) |  |  |
| 2 | 27 | ( | 17 | ) |  | 4 | ( | 33 | ) |  |  |  | 15 | ( | 14 | ) |  | 2 | ( | 6 | ) |  |  |
| 3 | 15 | ( | 17 | ) |  | 1 | ( | 8 | ) |  |  |  | 8 | ( | 7 | ) |  | 3 | ( | 10 | ) |  |  |
| 4 or more | 15 | ( | 9 | ) |  | 1 | ( | 8 | ) |  |  |  | 8 | ( | 7 | ) |  | 5 | ( | 16 | ) |  |  |
| Unknown/refused | 18 | ( | 9 | ) |  | 0 | ( | 0 | ) |  |  |  | 3 | ( | 3 | ) |  | 0 | ( | 0 | ) |  |  |
| Daily medications |  |  |  |  |  |  |  |  |  |  | 0.581 |  |  |  |  |  |  |  |  |  |  |  | 0.652 |
| 0 | 56 | ( | 35 | ) |  | 3 | ( | 25 | ) |  |  |  | 51 | ( | 47 | ) |  | 12 | ( | 39 | ) |  |  |
| 1 | 27 | ( | 17 | ) |  | 3 | ( | 25 | ) |  |  |  | 23 | ( | 21 | ) |  | 9 | ( | 29 | ) |  |  |
| 2 or more | 57 | ( | 36 | ) |  | 6 | ( | 50 | ) |  |  |  | 31 | ( | 29 | ) |  | 10 | ( | 32 | ) |  |  |
| Health behaviors |  |  |  |  |  |  |  |  |  |  |  |  |  |  |  |  |  |  |  |  |  |  |  |
| Smoking |  |  |  |  |  |  |  |  |  |  | 0.021 |  |  |  |  |  |  |  |  |  |  |  | 0.991 |
| Not current smoker | 137 | ( | 87 | ) |  | 7 | ( | 58 | ) |  |  |  | 87 | ( | 81 | ) |  | 25 | ( | 81 | ) |  |  |
| Smoke tobacco products | 21 | ( | 13 | ) |  | 5 | ( | 42 | ) |  |  |  | 21 | ( | 19 | ) |  | 6 | ( | 19 | ) |  |  |
| Influenza vaccination history in past 5 years |  |  |  |  |  |  |  |  |  |  | 0.212 |  |  |  |  |  |  |  |  |  |  |  | 0.687 |
| No vaccination history | 29 | ( | 18 | ) |  | 5 | ( | 42 | ) |  |  |  | 6 | ( | 6 | ) |  | 0 | ( | 0 | ) |  |  |
| 1 - 3 years of vaccination | 32 | ( | 20 | ) |  | 2 | ( | 17 | ) |  |  |  | 11 | ( | 10 | ) |  | 3 | ( | 10 | ) |  |  |
| 4 or more years of vaccination | 97 | ( | 61 | ) |  | 5 | ( | 42 | ) |  |  |  | 89 | ( | 82 | ) |  | 28 | ( | 90 | ) |  |  |
| Infection Characteristics |  |  |  |  |  |  |  |  |  |  |  |  |  |  |  |  |  |  |  |  |  |  |  |
| Symptomatic COVID-19 |  |  |  |  |  |  |  |  |  |  | 1.000 |  |  |  |  |  |  |  |  |  |  |  |  |
| COVID-19: CLI at RT-PCR-detection or onset <14 days of detection | 133 | ( | 84 | ) |  | 10 | ( | 83 | ) |  |  |  |  |  |  |  |  |  |  |  |  |  |  |
| COVID-19: CLI onset 2-14 day after RT-PCR-detection from weekly nasal swab | 13 | ( | 8 | ) |  | 1 | ( | 8 | ) |  |  |  |  |  |  |  |  |  |  |  |  |  |  |
| No CLI but other symptoms | 0 | ( | 0 | ) |  | 0 | ( | 0 | ) |  |  |  |  |  |  |  |  |  |  |  |  |  |  |
| Asymptomatic +/- 14-days of RT-PCR-detection from weekly nasal swab | 12 | ( | 8 | ) |  | 1 | ( | 8 | ) |  |  |  |  |  |  |  |  |  |  |  |  |  |  |
| Duration of RT-PCR-positive |  |  |  |  |  |  |  |  |  |  | 0.143 |  |  |  |  |  |  |  |  |  |  |  |  |
| 1 week | 33 | ( | 21 | ) |  | 5 | ( | 42 | ) |  |  |  |  |  |  |  |  |  |  |  |  |  |  |
| 2 or more weeks | 125 | ( | 79 | ) |  | 8 |  | 67 | ) |  |  |  |  |  |  |  |  |  |  |  |  |  |  |
| Febrile COVID-19 |  |  |  |  |  |  |  |  |  |  | 0.764 |  |  |  |  |  |  |  |  |  |  |  |  |
| Not febrile | 56 | ( | 35 | ) |  | 5 | ( | 42 | ) |  |  |  |  |  |  |  |  |  |  |  |  |  |  |
| Febrile | 95 | ( | 60 | ) |  | 7 | ( | 58 | ) |  |  |  |  |  |  |  |  |  |  |  |  |  |  |
| Unknown | 6 | ( | 4 | ) |  | 0 | ( | 0 | ) |  |  |  |  |  |  |  |  |  |  |  |  |  |  |
| Medically attended COVID-19 |  |  |  |  |  |  |  |  |  |  | 0.512 |  |  |  |  |  |  |  |  |  |  |  |  |
| No medical visit | 107 | ( | 68 | ) |  | 10 | ( | 83 | ) |  |  |  |  |  |  |  |  |  |  |  |  |  |  |
| One or more medical visits | 45 | ( | 28 | ) |  | 2 | ( | 17 | ) |  |  |  |  |  |  |  |  |  |  |  |  |  |  |
| Unknown | 6 | ( | 4 | ) |  | 0 | ( | 0 | ) |  |  |  |  |  |  |  |  |  |  |  |  |  |  |
| Emergency department visit |  |  |  |  |  |  |  |  |  |  | 1.000 |  |  |  |  |  |  |  |  |  |  |  |  |
| No | 143 | ( | 91 | ) |  | 12 | ( | 100 | ) |  |  |  |  |  |  |  |  |  |  |  |  |  |  |
| Yes | 9 | ( | 6 | ) |  | 0 | ( | 0 | ) |  |  |  |  |  |  |  |  |  |  |  |  |  |  |
| Unknown | 6 | ( | 4 | ) |  | 0 | ( | 0 | ) |  |  |  |  |  |  |  |  |  |  |  |  |  |  |
| Hospitalization |  |  |  |  |  |  |  |  |  |  | 1.000 |  |  |  |  |  |  |  |  |  |  |  |  |
| No | 150 | ( | 95 | ) |  | 12 | ( | 100 | ) |  |  |  |  |  |  |  |  |  |  |  |  |  |  |
| Yes | 2 | ( | 1 | ) |  | 0 | ( | 0 | ) |  |  |  |  |  |  |  |  |  |  |  |  |  |  |
| Unknown | 6 | ( | 4 | ) |  | 0 | ( | 0 | ) |  |  |  |  |  |  |  |  |  |  |  |  |  |  |
| Vaccination Characteristics |  |  |  |  |  |  |  |  |  |  |  |  |  |  |  |  |  |  |  |  |  |  |  |
| COVID-19 vaccine products |  |  |  |  |  |  |  |  |  |  |  |  |  |  |  |  |  |  |  |  |  |  | 0.112 |
| Pfizer-BioNTech |  |  |  |  |  |  |  |  |  |  |  |  | 75 | ( | 69 | ) |  | 26 | ( | 84 | ) |  |  |
| Moderna |  |  |  |  |  |  |  |  |  |  |  |  | 33 | ( | 31 | ) |  | 5 | ( | 16 | ) |  |  |
| Analgesic intake prior to dose 1 |  |  |  |  |  |  |  |  |  |  |  |  |  |  |  |  |  |  |  |  |  |  | 0.907 |
| No |  |  |  |  |  |  |  |  |  |  |  |  | 92 | ( | 85 | ) |  | 28 | ( | 90 | ) |  |  |
| Yes |  |  |  |  |  |  |  |  |  |  |  |  | 10 | ( | 9 | ) |  | 2 | ( | 6 | ) |  |  |
| Missing |  |  |  |  |  |  |  |  |  |  |  |  | 6 | ( | 6 | ) |  | 1 | ( | 3 | ) |  |  |
| Analgesic intake after dose 1 |  |  |  |  |  |  |  |  |  |  |  |  |  |  |  |  |  |  |  |  |  |  | 0.364 |
| No |  |  |  |  |  |  |  |  |  |  |  |  | 77 | ( | 71 | ) |  | 19 | ( | 61 | ) |  |  |
| Yes |  |  |  |  |  |  |  |  |  |  |  |  | 25 | ( | 23 | ) |  | 11 | ( | 35 | ) |  |  |
| Missing |  |  |  |  |  |  |  |  |  |  |  |  | 6 | ( | 6 | ) |  | 1 | ( | 3 | ) |  |  |
| Analgesic intake prior to dose 2 |  |  |  |  |  |  |  |  |  |  |  |  |  |  |  |  |  |  |  |  |  |  | 0.924 |
| No |  |  |  |  |  |  |  |  |  |  |  |  | 84 | ( | 78 | ) |  | 26 | ( | 84 | ) |  |  |
| Yes |  |  |  |  |  |  |  |  |  |  |  |  | 18 | ( | 17 | ) |  | 4 | ( | 13 | ) |  |  |
| Missing |  |  |  |  |  |  |  |  |  |  |  |  | 6 | ( | 6 | ) |  | 1 | ( | 3 | ) |  |  |
| Analgesic intake after dose 2 |  |  |  |  |  |  |  |  |  |  |  |  |  |  |  |  |  |  |  |  |  |  | 0.842 |
| No |  |  |  |  |  |  |  |  |  |  |  |  | 55 | ( | 51 | ) |  | 17 | ( | 55 | ) |  |  |
| Yes |  |  |  |  |  |  |  |  |  |  |  |  | 47 | ( | 44 | ) |  | 13 | ( | 42 | ) |  |  |
| Missing |  |  |  |  |  |  |  |  |  |  |  |  | 6 | ( | 6 | ) |  | 1 | ( | 3 | ) |  |  |
| Febrile after dose 1 |  |  |  |  |  |  |  |  |  |  |  |  |  |  |  |  |  |  |  |  |  |  | 0.716 |
| No |  |  |  |  |  |  |  |  |  |  |  |  | 93 | ( | 86 | ) |  | 29 | ( | 94 | ) |  |  |
| Yes |  |  |  |  |  |  |  |  |  |  |  |  | 9 | ( | 8 | ) |  | 1 | ( | 3 | ) |  |  |
| Missing |  |  |  |  |  |  |  |  |  |  |  |  | 6 | ( | 6 | ) |  | 1 | ( | 3 | ) |  |  |
| Febrile after dose 2 |  |  |  |  |  |  |  |  |  |  |  |  |  |  |  |  |  |  |  |  |  |  | 0.167 |
| No |  |  |  |  |  |  |  |  |  |  |  |  | 56 | ( | 52 | ) |  | 22 | ( | 71 | ) |  |  |
| Yes |  |  |  |  |  |  |  |  |  |  |  |  | 46 | ( | 43 | ) |  | 8 | ( | 26 | ) |  |  |
| Missing |  |  |  |  |  |  |  |  |  |  |  |  | 6 | ( | 6 | ) |  | 1 | ( | 3 | ) |  |  |

| **Supplementary Table 2. Geometric mean titers following SARS-CoV-2 infection and vaccination with messenger RNA (mRNA) COVID-19 vaccines** | | | | | | | | | | | | | | | | | | | | |
| --- | --- | --- | --- | --- | --- | --- | --- | --- | --- | --- | --- | --- | --- | --- | --- | --- | --- | --- | --- | --- |
|  | **Response to infection** | | | | |  | **Response to vaccination** | | | | |  | **Response to vaccination after infection** | | | | | | | |
|  |  |  | **N= 170** |  |  |  |  |  | **N= 139** |  |  |  |  |  |  | **N=50** |  |  |  |  |
|  | **All** | |  | **Nab positive only** | |  | **Post Dose 1** | |  | **Post Dose 2** | |  | **Post Infection** | |  | **Post Dose 1** | |  | **Post Dose 2** | |
|  | **N=170** |  |  | **N=158** |  |  | **N=139** |  |  | **N=139** |  |  | **N=50** |  |  | **N=42** |  |  | **N=40** |  |
|  | GMT | p-value |  | GMT | p-value |  | GMT | p-value |  | GMT | p-value |  | GMT | p-value |  | GMT | p-value |  | GMT | p-value |
| All participants (row %) | 1003 |  |  | 1351 |  |  | 201 |  |  | 2777 |  |  | 1438 |  |  | 21655 |  |  | 15863 |  |
| Socio-demographic characteristics |  |  |  |  |  |  |  |  |  |  |  |  |  |  |  |  |  |  |  |  |
| Cohort location |  | 0.083 |  |  | 0.269 |  |  | 0.006 |  |  | 0.009 |  |  | 0.800 |  |  | 0.178 |  |  | 0.065 |
| Phoenix, AZ | 1659 |  |  | 2093 |  |  |  |  |  |  |  |  | 2533 |  |  | 40018 |  |  |  |  |
| Tucson, AZ | 892 |  |  | 1280 |  |  | 775 |  |  | 4839 |  |  | 1056 |  |  | 32334 |  |  | 27666 |  |
| Other, AZ | 3083 |  |  | 3083 |  |  |  |  |  |  |  |  |  |  |  |  |  |  |  |  |
| Miami, FL | 338 |  |  | 807 |  |  | 760 |  |  | 7203 |  |  |  |  |  |  |  |  |  |  |
| Duluth, MN | 1390 |  |  | 1581 |  |  | 145 |  |  | 2487 |  |  | 1395 |  |  | 17145 |  |  | 14689 |  |
| Portland, OR | 471 |  |  | 471 |  |  | 293 |  |  | 2729 |  |  |  |  |  |  |  |  |  |  |
| Temple, TX | 1007 |  |  | 1396 |  |  | 88 |  |  | 1535 |  |  | 2966 |  |  | 18414 |  |  | 13888 |  |
| Salt Lake City, UT | 1405 |  |  | 1405 |  |  | 358 |  |  | 11998 |  |  | 1453 |  |  |  |  |  | 8311 |  |
| Sex |  | 0.138 |  |  | 0.103 |  |  | 0.164 |  |  | 0.161 |  |  | 0.845 |  |  | 0.047 |  |  | 0.130 |
| Female | 1212 |  |  | 1614 |  |  | 169 |  |  | 2483 |  |  | 1377 |  |  | 17576 |  |  | 13526 |  |
| Male | 803 |  |  | 1093 |  |  | 240 |  |  | 3112 |  |  | 1526 |  |  | 31528 |  |  | 19681 |  |
| Age (Years) |  | 0.369 |  |  | 0.045 |  |  | 0.045 |  |  | 0.155 |  |  | 0.795 |  |  | 0.005 |  |  | 0.013 |
| 18-49 | 924 |  |  | 1172 |  |  | 241 |  |  | 3026 |  |  | 1359 |  |  | 15627 |  |  | 13441 |  |
| ≥50 | 1223 |  |  | 1932 |  |  | 139 |  |  | 2322 |  |  | 1589 |  |  | 36793 |  |  | 26075 |  |
| Race |  |  |  |  |  |  |  |  |  |  |  |  |  |  |  |  |  |  |  |  |
| White |  |  |  |  |  |  |  |  |  |  |  |  |  |  |  |  |  |  |  |  |
| Other |  |  |  |  |  |  |  |  |  |  |  |  |  |  |  |  |  |  |  |  |
| Ethnicity |  |  |  |  |  |  |  |  |  |  |  |  |  |  |  |  |  |  |  |  |
| Hispanic/Latinx |  |  |  |  |  |  |  |  |  |  |  |  |  |  |  |  |  |  |  |  |
| Other |  |  |  |  |  |  |  |  |  |  |  |  |  |  |  |  |  |  |  |  |
| Occupation |  |  |  |  |  |  |  |  |  |  |  |  |  |  |  |  |  |  |  |  |
| Primary HCP |  |  |  |  |  |  |  |  |  |  |  |  |  |  |  |  |  |  |  |  |
| Nurses and other allied HCP |  |  |  |  |  |  |  |  |  |  |  |  |  |  |  |  |  |  |  |  |
| First Responders |  |  |  |  |  |  |  |  |  |  |  |  |  |  |  |  |  |  |  |  |
| Essential and other frontline |  |  |  |  |  |  |  |  |  |  |  |  |  |  |  |  |  |  |  |  |
| Health status |  |  |  |  |  |  |  |  |  |  |  |  |  |  |  |  |  |  |  |  |
| BMI |  | 0.480 |  |  | 0.242 |  |  | 0.095 |  |  | 0.019 |  |  | 0.187 |  |  | 0.250 |  |  | 0.230 |
| Underweight | 398 |  |  | 398 |  |  | 446 |  |  | 3409 |  |  |  |  |  |  |  |  |  |  |
| Normal weight | 837 |  |  | 1068 |  |  | 142 |  |  | 1904 |  |  | 697 |  |  | 14935 |  |  | 11131 |  |
| Overweight | 944 |  |  | 1334 |  |  | 289 |  |  | 3071 |  |  | 1827 |  |  | 21585 |  |  | 18508 |  |
| Obese | 1414 |  |  | 1916 |  |  | 165 |  |  | 3636 |  |  | 2094 |  |  | 28847 |  |  | 16510 |  |
| Missing |  |  |  |  |  |  |  |  |  |  |  |  |  |  |  |  |  |  |  |  |
| Self-Rated Health |  | 0.103 |  |  | 0.005 |  |  | 0.271 |  |  | 0.974 |  |  | 0.525 |  |  | 0.089 |  |  | 0.639 |
| Excellent/Very good | 819 |  |  | 1011 |  |  | 220 |  |  | 2782 |  |  | 1260 |  |  | 17315 |  |  | 16428 |  |
| Good/Fair/Poor | 1283 |  |  | 1944 |  |  | 162 |  |  | 2766 |  |  | 1726 |  |  | 28387 |  |  | 14466 |  |
| Chronic Condition |  | 0.286 |  |  | 0.130 |  |  | 0.033 |  |  | 0.393 |  |  | 0.969 |  |  | 0.016 |  |  | 0.021 |
| None | 925 |  |  | 1231 |  |  | 247 |  |  | 2927 |  |  | 1428 |  |  | 17145 |  |  | 13143 |  |
| 1 or more | 1306 |  |  | 1832 |  |  | 136 |  |  | 2507 |  |  | 1460 |  |  | 36456 |  |  | 23446 |  |
| Chronic Condition and >= 1 daily medication |  | 0.398 |  |  | 0.176 |  |  | 0.156 |  |  | 0.484 |  |  | 0.910 |  |  | 0.011 |  |  | 0.018 |
| No | 943 |  |  | 1247 |  |  | 224 |  |  | 2885 |  |  | 1409 |  |  | 17071 |  |  | 13165 |  |
| 1 or more | 1255 |  |  | 1809 |  |  | 150 |  |  | 2500 |  |  | 1508 |  |  | 39246 |  |  | 24508 |  |
| Daily medications |  | 0.480 |  |  | 0.135 |  |  | 0.526 |  |  | 0.160 |  |  | 0.444 |  |  | 0.161 |  |  | 0.093 |
| 0 | 742 |  |  | 901 |  |  | 226 |  |  | 2882 |  |  | 908 |  |  | 12506 |  |  | 11376 |  |
| 1 | 787 |  |  | 1184 |  |  | 160 |  |  | 2154 |  |  | 2484 |  |  | 28312 |  |  | 20095 |  |
| 2 | 902 |  |  | 1586 |  |  | 288 |  |  | 3578 |  |  | 905 |  |  | 22034 |  |  | 22224 |  |
| 3 | 1456 |  |  | 1938 |  |  | 141 |  |  | 2091 |  |  | 2879 |  |  | 34182 |  |  | 25563 |  |
| 4 or more | 1532 |  |  | 2046 |  |  | 153 |  |  | 4029 |  |  | 1732 |  |  | 29018 |  |  | 13752 |  |
| Unknown/refused |  |  |  |  |  |  |  |  |  |  |  |  |  |  |  |  |  |  |  |  |
| Daily medications |  | 0.331 |  |  | 0.035 |  |  | 0.556 |  |  | 0.187 |  |  | 0.417 |  |  | 0.055 |  |  | 0.051 |
| 0 | 742 |  |  | 901 |  |  | 226 |  |  | 2882 |  |  | 908 |  |  | 12506 |  |  | 11376 |  |
| 1 | 787 |  |  | 1184 |  |  | 160 |  |  | 2154 |  |  | 2484 |  |  | 28312 |  |  | 20095 |  |
| 2 or more | 1165 |  |  | 1788 |  |  | 194 |  |  | 6217 |  |  | 1368 |  |  | 26120 |  |  | 20322 |  |
| Health behaviors |  |  |  |  |  |  |  |  |  |  |  |  |  |  |  |  |  |  |  |  |
| Smoking |  | 0.350 |  |  | 0.667 |  |  | 0.932 |  |  | 0.916 |  |  | 0.800 |  |  | 0.946 |  |  | 0.474 |
| Not current smoker | 1076 |  |  | 1320 |  |  | 203 |  |  | 2765 |  |  | 1373 |  |  | 21531 |  |  | 15074 |  |
| Smoke tobacco products | 680 |  |  | 1574 |  |  | 197 |  |  | 2825 |  |  | 1696 |  |  | 22113 |  |  | 19455 |  |
| Influenza vaccination history in past 5 years |  | 0.108 |  |  | 0.346 |  |  | 0.078 |  |  | 0.076 |  |  | 0.265 |  |  | 0.878 |  |  | 0.767 |
| No vaccination history | 621 |  |  | 1124 |  |  | 708 |  |  | 9472 |  |  | 3836 |  |  | 24612 |  |  | 16720 |  |
| 1 - 3 years of vaccination | 818 |  |  | 1048 |  |  | 381 |  |  | 2711 |  |  | 587 |  |  | 17356 |  |  | 19466 |  |
| 4 or more years of vaccination | 1250 |  |  | 1540 |  |  | 183 |  |  | 2702 |  |  | 1526 |  |  | 22036 |  |  | 15216 |  |
| Infection Characteristics |  |  |  |  |  |  |  |  |  |  |  |  |  |  |  |  |  |  |  |  |
| Symptomatic COVID-19 |  | 0.093 |  |  | 0.018 |  |  |  |  |  |  |  |  | 0.444 |  |  | 0.077 |  |  | 0.561 |
| COVID-19: CLI at PCR-detection or onset <14 days of detection | 882 |  |  | 1173 |  |  |  |  |  |  |  |  | 1341 |  |  | 18426 |  |  | 14942 |  |
| COVID-19: CLI onset 2-14 day after PCR-detection from weekly nasal swab | 1820 |  |  | 2575 |  |  |  |  |  |  |  |  | 3160 |  |  | 33611 |  |  | 20724 |  |
| No CLI but other symptoms |  |  |  |  |  |  |  |  |  |  |  |  |  |  |  |  |  |  |  |  |
| Asymptomatic +/- 14-days of PCR-detection from weekly nasal swab | 2173 |  |  | 3212 |  |  |  |  |  |  |  |  | 888 |  |  | 53102 |  |  | 21443 |  |
| Duration of PCR-positive |  | 0.717 |  |  | 0.524 |  |  |  |  |  |  |  |  | 0.934 |  |  | 0.589 |  |  | 0.134 |
| 1 week | 899 |  |  | 1600 |  |  |  |  |  |  |  |  | 1540 |  |  | 18127 |  |  | 26368 |  |
| 2 or more weeks | 1036 |  |  | 1292 |  |  |  |  |  |  |  |  | 1417 |  |  | 22580 |  |  | 14502 |  |
| Febrile COVID-19 |  | 0.801 |  |  | 0.577 |  |  |  |  |  |  |  |  | 0.821 |  |  | 0.166 |  |  | 0.301 |
| Not febrile | 1035 |  |  | 1472 |  |  |  |  |  |  |  |  | 1334 |  |  | 28406 |  |  | 18979 |  |
| Febrile | 959 |  |  | 1276 |  |  |  |  |  |  |  |  | 1500 |  |  | 18624 |  |  | 14551 |  |
| Unknown |  |  |  |  |  |  |  |  |  |  |  |  |  |  |  |  |  |  |  |  |
| Medically attended COVID-19 |  | 0.649 |  |  | 0.876 |  |  |  |  |  |  |  |  | 0.128 |  |  | 0.497 |  |  | 0.581 |
| No medical visit | 974 |  |  | 1401 |  |  |  |  |  |  |  |  | 1188 |  |  | 23082 |  |  | 16583 |  |
| One or more medical visits | 1122 |  |  | 1342 |  |  |  |  |  |  |  |  | 2475 |  |  | 18460 |  |  | 13615 |  |
| Unknown |  |  |  |  |  |  |  |  |  |  |  |  |  |  |  |  |  |  |  |  |
| Emergency department visit |  | 0.505 |  |  | 0.878 |  |  |  |  |  |  |  |  | 0.120 |  |  | 0.830 |  |  | - |
| No | 991 |  |  | 1376 |  |  |  |  |  |  |  |  | 1367 |  |  | 21258 |  |  | 16332 |  |
| Yes | 1513 |  |  | 1513 |  |  |  |  |  |  |  |  | 4826 |  |  | 31341 |  |  | 5093 |  |
| Unknown |  |  |  |  |  |  |  |  |  |  |  |  |  |  |  |  |  |  |  |  |
| Hospitalization |  | 0.935 |  |  | 0.846 |  |  |  |  |  |  |  |  |  |  |  |  |  |  |  |
| No | 1017 |  |  | 1393 |  |  |  |  |  |  |  |  |  |  |  |  |  |  |  |  |
| Yes | 813 |  |  | 813 |  |  |  |  |  |  |  |  |  |  |  |  |  |  |  |  |
| Unknown |  |  |  |  |  |  |  |  |  |  |  |  |  |  |  |  |  |  |  |  |
| Vaccination Characteristics |  |  |  |  |  |  |  |  |  |  |  |  |  |  |  |  |  |  |  |  |
| COVID-19 vaccine products |  |  |  |  |  |  |  | 0.001 |  |  | 0.000 |  |  | 0.853 |  |  | 0.008 |  |  | 0.154 |
| Pfizer-BioNTech |  |  |  |  |  |  | 156 |  |  | 2309 |  |  | 1488 |  |  | 18243 |  |  | 14635 |  |
| Moderna |  |  |  |  |  |  | 401 |  |  | 4539 |  |  | 1274 |  |  | 51029 |  |  | 25047 |  |
| Analgesic intake prior to dose 1 |  |  |  |  |  |  |  | 0.973 |  |  | 0.842 |  |  | 0.968 |  |  | 0.066 |  |  | 0.017 |
| No |  |  |  |  |  |  | 197 |  |  | 2752 |  |  | 1701 |  |  | 18716 |  |  | 14105 |  |
| Yes |  |  |  |  |  |  | 195 |  |  | 2623 |  |  | 1648 |  |  | 35853 |  |  | 28490 |  |
| Missing |  |  |  |  |  |  |  |  |  |  |  |  |  |  |  |  |  |  |  |  |
| Analgesic intake after dose 1 |  |  |  |  |  |  |  | 0.424 |  |  | 0.218 |  |  | 0.096 |  |  | 0.005 |  |  | 0.028 |
| No |  |  |  |  |  |  | 210 |  |  | 2593 |  |  | 1123 |  |  | 14005 |  |  | 12536 |  |
| Yes |  |  |  |  |  |  | 165 |  |  | 3194 |  |  | 2546 |  |  | 31780 |  |  | 22233 |  |
| Missing |  |  |  |  |  |  |  |  |  |  |  |  |  |  |  |  |  |  |  |  |
| Analgesic intake prior to dose 2 |  |  |  |  |  |  |  | 0.876 |  |  | 0.219 |  |  | 0.397 |  |  | 0.841 |  |  | 0.565 |
| No |  |  |  |  |  |  | 195 |  |  | 2630 |  |  | 1949 |  |  | 20707 |  |  | 15352 |  |
| Yes |  |  |  |  |  |  | 205 |  |  | 3399 |  |  | 1206 |  |  | 22086 |  |  | 18200 |  |
| Missing |  |  |  |  |  |  |  |  |  |  |  |  |  |  |  |  |  |  |  |  |
| Analgesic intake after dose 2 |  |  |  |  |  |  |  | 0.274 |  |  | 0.031 |  |  | 0.225 |  |  | 0.031 |  |  | 0.171 |
| No |  |  |  |  |  |  | 174 |  |  | 2351 |  |  | 1265 |  |  | 15577 |  |  | 13976 |  |
| Yes |  |  |  |  |  |  | 230 |  |  | 3312 |  |  | 2323 |  |  | 29554 |  |  | 19836 |  |
| Missing |  |  |  |  |  |  |  |  |  |  |  |  |  |  |  |  |  |  |  |  |
| Febrile after dose 1 |  |  |  |  |  |  |  | 0.104 |  |  |  |  |  |  |  |  | 0.045 |  |  |  |
| No |  |  |  |  |  |  | 183 |  |  |  |  |  |  |  |  | 18198 |  |  |  |  |
| Yes |  |  |  |  |  |  | 400 |  |  |  |  |  |  |  |  | 33968 |  |  |  |  |
| Missing |  |  |  |  |  |  |  |  |  |  |  |  |  |  |  |  |  |  |  |  |
| Febrile after dose 2 |  |  |  |  |  |  |  |  |  |  | 0.004 |  |  |  |  |  |  |  |  | 0.014 |
| No |  |  |  |  |  |  |  |  |  | 2236 |  |  |  |  |  |  |  |  | 13222 |  |
| Yes |  |  |  |  |  |  |  |  |  | 3587 |  |  |  |  |  |  |  |  | 25554 |  |
| Missing |  |  |  |  |  |  |  |  |  |  |  |  |  |  |  |  |  |  |  |  |

| **Supplementary Table S3. Detection and titer of neutralizing antibodies (nAb) to SARS-CoV-2 vaccination with messenger RNA (mRNA) COVID-19 vaccines among previously infected participants with a complete blood draw series** | | | | | | | | | | | | | | | | | | | | | | | | |
| --- | --- | --- | --- | --- | --- | --- | --- | --- | --- | --- | --- | --- | --- | --- | --- | --- | --- | --- | --- | --- | --- | --- | --- | --- |
|  | Participants | | | |  | Detection of SARS-CoV-2 nAb | | | | | | | | |  | Adjusted Quantity of nAb | | | |  | Adjusted Fold-Change in nAb Response † | | | |
|  |  |  |  |  |  | None | | | |  | Detected nAb | | | |  |  |  |  |  |  |  |  |  |  |
|  | N | ( | Col % | ) |  | N | ( | Row % | ) |  | N | ( | Row % | ) |  | aGMT | ( | 95% CI | ) |  | GMR | ( | 95% CI | ) |
| Response to mRNA vaccination following natural infection with SARS-CoV-2 |  |  |  |  |  |  |  |  |  |  |  |  |  |  |  |  |  |  |  |  |  |  |  |  |
| Either mRNA vaccines |  |  |  |  |  |  |  |  |  |  |  |  |  |  |  |  |  |  |  |  |  |  |  |  |
| After infection, median (IQR) = 28 (24-30) days | 32 | ( | 100 | ) |  | 0 | ( | 0 |  |  | 32 | ( | 100 | ) |  | 1641 | ( | 1167-2306 | ) |  |  |  |  |  |
| After dose-1, median (IQR) = 15 (14-20) days | 32 | ( | 100 | ) |  | 0 | ( | 0 | ) |  | 32 | ( | 100 | ) |  | 18492 | ( | 13159-25988 | ) |  | 9.7 | ( | 6.0-15.7 | ) |
| After dose-2, median (IQR) = 16 (14-21) days | 32 | ( | 100 | ) |  | 0 | ( | 0 | ) |  | 32 | ( | 100 | ) |  | 15970 | ( | 11364-22443 | ) |  | 0.9 | ( | 0.5-1.4 | ) |
| Abbreviations: Neutralizing antibodies (nAb); Messenger RNA (mRNA) |  |  |  |  |  |  |  |  |  |  |  |  |  |  |  |  |  |  |  |  |  |  |  |  |
| † Geometric mean ratio is fold-change in nAb from dose-1 to dose-2 from repeated measures ANOVA. | | | | | | | | | | | | | | | | | | | | | | | | |

| **Supplementary Table 4. Participant socio-demographic and health characteristics, SARS-CoV-2 infection characteristics, and vaccination information by vaccine product received** | | | | | | | | | | | | | | | | | | | | | | | |
| --- | --- | --- | --- | --- | --- | --- | --- | --- | --- | --- | --- | --- | --- | --- | --- | --- | --- | --- | --- | --- | --- | --- | --- |
|  | **Response to vaccination among those uninfected** | | | | | | | | | | |  | **Response to vaccination after PCR confirmed infection** | | | | | | | | | | |
|  | **BNT162b2** | | | |  | **mRNA-1273** | | | |  |  |  | **BNT162b2** | | | |  | **mRNA-1273** | | | |  |  |
|  | N | ( | Col. % | ) |  | N | ( | Col. % | ) |  | p-value |  | N | ( | Col. % | ) |  | N | ( | Col. % | ) |  | p-value |
| All participants (row %) | 101 |  |  |  |  | 38 |  |  |  |  |  |  | 39 |  |  |  |  | 11 |  |  |  |  |  |
| Socio-demographic characteristics |  |  |  |  |  |  |  |  |  |  |  |  |  |  |  |  |  |  |  |  |  |  |  |
| Cohort location |  |  |  |  |  |  |  |  |  |  | <.001 |  |  |  |  |  |  |  |  |  |  |  | <.001 |
| Phoenix, AZ | 0 | ( | 0 | ) |  | 0 | ( | 0 | ) |  |  |  | 3 | ( | 8 | ) |  | 0 | ( | 0 | ) |  |  |
| Tucson, AZ | 1 | ( | 1 | ) |  | 2 | ( | 5 | ) |  |  |  | 4 | ( | 10 | ) |  | 11 | ( | 100 | ) |  |  |
| Other, AZ | 0 | ( | 0 | ) |  | 0 | ( | 0 | ) |  |  |  | 0 | ( | 0 | ) |  | 0 | ( | 0 | ) |  |  |
| Miami, FL | 1 | ( | 1 | ) |  | 6 | ( | 16 | ) |  |  |  | 0 | ( | 0 | ) |  | 0 | ( | 0 | ) |  |  |
| Duluth, MN | 71 | ( | 70 | ) |  | 14 | ( | 37 | ) |  |  |  | 24 | ( | 62 | ) |  | 0 | ( | 0 | ) |  |  |
| Portland, OR | 27 | ( | 27 | ) |  | 13 | ( | 34 | ) |  |  |  | 0 | ( | 0 | ) |  | 0 | ( | 0 | ) |  |  |
| Temple, TX | 0 | ( | 0 | ) |  | 2 | ( | 5 | ) |  |  |  | 5 | ( | 13 | ) |  | 0 | ( | 0 | ) |  |  |
| Salt Lake City, UT | 1 | ( | 1 | ) |  | 1 | ( | 3 | ) |  |  |  | 3 | ( | 8 | ) |  | 0 | ( | 0 | ) |  |  |
| Sex |  |  |  |  |  |  |  |  |  |  | 0.023 |  |  |  |  |  |  |  |  |  |  |  | <.001 |
| Female | 57 | ( | 56 | ) |  | 13 | ( | 34 | ) |  |  |  | 28 | ( | 72 | ) |  | 1 | ( | 9 | ) |  |  |
| Male | 44 | ( | 44 | ) |  | 25 | ( | 66 | ) |  |  |  | 11 | ( | 28 | ) |  | 10 | ( | 91 | ) |  |  |
| Age (Years) |  |  |  |  |  |  |  |  |  |  | 0.840 |  |  |  |  |  |  |  |  |  |  |  | 0.172 |
| 18-49 | 69 | ( | 68 | ) |  | 25 | ( | 66 | ) |  |  |  | 27 | ( | 69 | ) |  | 5 | ( | 45 | ) |  |  |
| ≥50 | 32 | ( | 32 | ) |  | 13 | ( | 34 | ) |  |  |  | 12 | ( | 31 | ) |  | 6 | ( | 55 | ) |  |  |
| Race |  |  |  |  |  |  |  |  |  |  | 0.736 |  |  |  |  |  |  |  |  |  |  |  | 0.206 |
| White | 93 | ( | 92 | ) |  | 34 | ( | 89 | ) |  |  |  | 37 | ( | 95 | ) |  | 9 | ( | 82 | ) |  |  |
| Other | 8 | ( | 8 | ) |  | 4 | ( | 11 | ) |  |  |  | 2 | ( | 5 | ) |  | 2 | ( | 18 | ) |  |  |
| Ethnicity |  |  |  |  |  |  |  |  |  |  | 0.575 |  |  |  |  |  |  |  |  |  |  |  | 0.004 |
| Hispanic/Latinx | 4 | ( | 4 | ) |  | 0 | ( | 0 | ) |  |  |  | 2 | ( | 5 | ) |  | 5 | ( | 45 | ) |  |  |
| Other | 97 | ( | 96 | ) |  | 38 | ( | 100 | ) |  |  |  | 37 | ( | 95 | ) |  | 6 | ( | 55 | ) |  |  |
| Marital status |  |  |  |  |  |  |  |  |  |  | 0.503 |  |  |  |  |  |  |  |  |  |  |  | 1.000 |
| Married | 76 | ( | 75 | ) |  | 31 | ( | 82 | ) |  |  |  | 26 | ( | 67 | ) |  | 7 | ( | 64 | ) |  |  |
| Other | 25 | ( | 25 | ) |  | 7 | ( | 18 | ) |  |  |  | 13 | ( | 33 | ) |  | 4 | ( | 36 | ) |  |  |
| Occupation |  |  |  |  |  |  |  |  |  |  |  |  |  |  |  |  |  |  |  |  |  |  |  |
| Occupation ¶ |  |  |  |  |  |  |  |  |  |  | <.001 |  |  |  |  |  |  |  |  |  |  |  | <.001 |
| Primary HCP | 37 | ( | 37 | ) |  | 2 | ( | 5 | ) |  |  |  | 6 | ( | 15 | ) |  | 0 | ( | 0 | ) |  |  |
| Nurses and other allied HCP | 60 | ( | 59 | ) |  | 12 | ( | 32 | ) |  |  |  | 28 | ( | 72 | ) |  | 1 | ( | 9 | ) |  |  |
| First Responders | 4 | ( | 4 | ) |  | 23 | ( | 61 | ) |  |  |  | 3 | ( | 8 | ) |  | 9 | ( | 82 | ) |  |  |
| Essential and other frontline | 0 | ( | 0 | ) |  | 1 | ( | 3 | ) |  |  |  | 2 | ( | 5 | ) |  | 1 | ( | 9 | ) |  |  |
| Health status |  |  |  |  |  |  |  |  |  |  |  |  |  |  |  |  |  |  |  |  |  |  |  |
| BMI |  |  |  |  |  |  |  |  |  |  | 0.129 |  |  |  |  |  |  |  |  |  |  |  | 0.144 |
| Underweight | 0 | ( | 0 | ) |  | 1 | ( | 3 | ) |  |  |  | 0 | ( | 0 | ) |  | 0 | ( | 0 | ) |  |  |
| Normal weight | 33 | ( | 33 | ) |  | 7 | ( | 18 | ) |  |  |  | 12 | ( | 31 | ) |  | 1 | ( | 9 | ) |  |  |
| Overweight | 38 | ( | 38 | ) |  | 18 | ( | 47 | ) |  |  |  | 15 | ( | 38 | ) |  | 8 | ( | 73 | ) |  |  |
| Obese | 26 | ( | 26 | ) |  | 12 | ( | 32 | ) |  |  |  | 11 | ( | 28 | ) |  | 2 | ( | 18 | ) |  |  |
| Missing | 4 | ( | 4 | ) |  | 0 | ( | 0 | ) |  |  |  | 1 | ( | 3 | ) |  | 0 | ( | 0 | ) |  |  |
| Self-Rated Health |  |  |  |  |  |  |  |  |  |  | 1.000 |  |  |  |  |  |  |  |  |  |  |  | 1.000 |
| Excellent/Very good | 72 | ( | 71 | ) |  | 27 | ( | 71 | ) |  |  |  | 23 | ( | 59 | ) |  | 6 | ( | 55 | ) |  |  |
| Good/Fair/Poor | 29 | ( | 29 | ) |  | 11 | ( | 29 | ) |  |  |  | 16 | ( | 41 | ) |  | 5 | ( | 45 | ) |  |  |
| Chronic Condition |  |  |  |  |  |  |  |  |  |  | 0.307 |  |  |  |  |  |  |  |  |  |  |  | 0.297 |
| None | 73 | ( | 72 | ) |  | 24 | ( | 63 | ) |  |  |  | 28 | ( | 72 | ) |  | 6 | ( | 55 | ) |  |  |
| 1 or more | 28 | ( | 28 | ) |  | 14 | ( | 37 | ) |  |  |  | 11 | ( | 28 | ) |  | 5 | ( | 45 | ) |  |  |
| Chronic Condition and >= 1 daily medication |  |  |  |  |  |  |  |  |  |  | 0.281 |  |  |  |  |  |  |  |  |  |  |  | 0.269 |
| No | 77 | ( | 76 | ) |  | 25 | ( | 66 | ) |  |  |  | 29 | ( | 74 | ) |  | 6 | ( | 55 | ) |  |  |
| 1 or more | 24 | ( | 24 | ) |  | 13 | ( | 34 | ) |  |  |  | 10 | ( | 26 | ) |  | 5 | ( | 45 | ) |  |  |
| Daily medications |  |  |  |  |  |  |  |  |  |  | 0.901 |  |  |  |  |  |  |  |  |  |  |  | 0.766 |
| 0 | 48 | ( | 48 | ) |  | 17 | ( | 45 | ) |  |  |  | 14 | ( | 36 | ) |  | 3 | ( | 27 | ) |  |  |
| 1 | 24 | ( | 24 | ) |  | 8 | ( | 21 | ) |  |  |  | 4 | ( | 10 | ) |  | 2 | ( | 18 | ) |  |  |
| 2 | 12 | ( | 12 | ) |  | 5 | ( | 13 | ) |  |  |  | 10 | ( | 26 | ) |  | 3 | ( | 27 | ) |  |  |
| 3 | 8 | ( | 8 | ) |  | 3 | ( | 8 | ) |  |  |  | 5 | ( | 13 | ) |  | 0 | ( | 0 | ) |  |  |
| 4 or more | 8 | ( | 8 | ) |  | 5 | ( | 13 | ) |  |  |  | 5 | ( | 13 | ) |  | 2 | ( | 18 | ) |  |  |
| Unknown/refused | 1 | ( | 1 | ) |  | 0 | ( | 0 | ) |  |  |  | 1 | ( | 3 | ) |  | 1 | ( | 9 | ) |  |  |
| Health behaviors |  |  |  |  |  |  |  |  |  |  |  |  |  |  |  |  |  |  |  |  |  |  |  |
| Smoking |  |  |  |  |  |  |  |  |  |  | 0.817 |  |  |  |  |  |  |  |  |  |  |  | 0.688 |
| Not current smoker | 80 | ( | 79 | ) |  | 31 | ( | 82 | ) |  |  |  | 31 | ( | 79 | ) |  | 8 | ( | 73 | ) |  |  |
| Smoke tobacco products | 21 | ( | 21 | ) |  | 7 | ( | 18 | ) |  |  |  | 8 | ( | 21 | ) |  | 3 | ( | 27 | ) |  |  |
| Influenza vaccination history in past 5 years |  |  |  |  |  |  |  |  |  |  | 0.031 |  |  |  |  |  |  |  |  |  |  |  | 0.016 |
| No vaccination history | 0 | ( | 0 | ) |  | 3 | ( | 8 | ) |  |  |  | 2 | ( | 5 | ) |  | 1 | ( | 9 | ) |  |  |
| 1 - 3 years of vaccination | 9 | ( | 9 | ) |  | 4 | ( | 11 | ) |  |  |  | 2 | ( | 5 | ) |  | 4 | ( | 36 | ) |  |  |
| 4 or more years of vaccination | 92 | ( | 91 | ) |  | 31 | ( | 82 | ) |  |  |  | 35 | ( | 90 | ) |  | 6 | ( | 55 | ) |  |  |
| Infection Characteristics |  |  |  |  |  |  |  |  |  |  |  |  |  |  |  |  |  |  |  |  |  |  |  |
| Specimen type at first PCR-positive |  |  |  |  |  |  |  |  |  |  |  |  |  |  |  |  |  |  |  |  |  |  | 0.498 |
| Nasal swab and saliva (CLI kit) |  |  |  |  |  |  |  |  |  |  |  |  | 16 | ( | 41 | ) |  | 3 | ( | 27 | ) |  |  |
| Weekly nasal swab |  |  |  |  |  |  |  |  |  |  |  |  | 23 | ( | 59 | ) |  | 8 | ( | 73 | ) |  |  |
| Unknown |  |  |  |  |  |  |  |  |  |  |  |  | 0 | ( | 0 | ) |  | 0 | ( | 0 | ) |  |  |
| Symptomatic COVID-19^‖‖^ |  |  |  |  |  |  |  |  |  |  |  |  |  |  |  |  |  |  |  |  |  |  | 0.688 |
| COVID-19: CLI at PCR-detection or onset <14 days of detection |  |  |  |  |  |  |  |  |  |  |  |  | 32 | ( | 82 | ) |  | 8 | ( | 73 | ) |  |  |
| COVID-19: CLI onset 2-14 day after PCR-detection from weekly nasal swab |  |  |  |  |  |  |  |  |  |  |  |  | 4 | ( | 10 | ) |  | 2 | ( | 18 | ) |  |  |
| No CLI but other symptoms |  |  |  |  |  |  |  |  |  |  |  |  | 0 | ( | 0 | ) |  | 0 | ( | 0 | ) |  |  |
| Asymptomatic +/- 14-days of PCR-detection from weekly nasal swab |  |  |  |  |  |  |  |  |  |  |  |  | 3 | ( | 8 | ) |  | 1 | ( | 9 | ) |  |  |
| Unknown |  |  |  |  |  |  |  |  |  |  |  |  | 0 | ( | 0 | ) |  | 0 | ( | 0 | ) |  |  |
| Febrile COVID-19 |  |  |  |  |  |  |  |  |  |  |  |  |  |  |  |  |  |  |  |  |  |  | 0.172 |
| Not febrile |  |  |  |  |  |  |  |  |  |  |  |  | 12 | ( | 31 | ) |  | 6 | ( | 55 | ) |  |  |
| Febrile |  |  |  |  |  |  |  |  |  |  |  |  | 27 | ( | 69 | ) |  | 5 | ( | 45 | ) |  |  |
| Unknown |  |  |  |  |  |  |  |  |  |  |  |  | 0 | ( | 0 | ) |  | 0 | ( | 0 | ) |  |  |
| Medically attended COVID-19 |  |  |  |  |  |  |  |  |  |  |  |  |  |  |  |  |  |  |  |  |  |  | 0.704 |
| No medical visit |  |  |  |  |  |  |  |  |  |  |  |  | 28 | ( | 72 | ) |  | 9 | ( | 82 | ) |  |  |
| One or more medical visits |  |  |  |  |  |  |  |  |  |  |  |  | 11 | ( | 28 | ) |  | 2 | ( | 18 | ) |  |  |
| Unknown |  |  |  |  |  |  |  |  |  |  |  |  | 0 | ( | 0 | ) |  | 0 | ( | 0 | ) |  |  |
| Emergency department visit |  |  |  |  |  |  |  |  |  |  |  |  |  |  |  |  |  |  |  |  |  |  | 0.395 |
| No |  |  |  |  |  |  |  |  |  |  |  |  | 38 | ( | 97 | ) |  | 10 | ( | 91 | ) |  |  |
| Yes |  |  |  |  |  |  |  |  |  |  |  |  | 1 | ( | 3 | ) |  | 1 | ( | 9 | ) |  |  |
| Unknown |  |  |  |  |  |  |  |  |  |  |  |  | 0 | ( | 0 | ) |  | 0 | ( | 0 | ) |  |  |
| Hospitalization |  |  |  |  |  |  |  |  |  |  |  |  |  |  |  |  |  |  |  |  |  |  | 0.22 |
| No |  |  |  |  |  |  |  |  |  |  |  |  | 39 | ( | 100 | ) |  | 10 | ( | 91 | ) |  |  |
| Yes |  |  |  |  |  |  |  |  |  |  |  |  | 0 | ( | 0 | ) |  | 1 | ( | 9 | ) |  |  |
| Unknown |  |  |  |  |  |  |  |  |  |  |  |  | 0 | ( | 0 | ) |  | 0 | ( | 0 | ) |  |  |
| Vaccination Characteristics |  |  |  |  |  |  |  |  |  |  |  |  |  |  |  |  |  |  |  |  |  |  |  |
| Vaccine doses received |  |  |  |  |  |  |  |  |  |  | - |  |  |  |  |  |  |  |  |  |  |  | - |
| One mRNA dose | 0 | ( | 0 | ) |  | 0 | ( | 0 | ) |  |  |  | 0 | ( | 0 | ) |  | 0 | ( | 0 | ) |  |  |
| Two mRNA doses | 101 | ( | 100 | ) |  | 38 | ( | 100 | ) |  |  |  | 39 | ( | 100 | ) |  | 11 | ( | 100 | ) |  |  |
| One J&J dose | 0 | ( | 0 | ) |  | 0 | ( | 0 | ) |  |  |  | 0 | ( | 0 | ) |  | 0 | ( | 0 | ) |  |  |
| None | 0 | ( | 0 | ) |  | 0 | ( | 0 | ) |  |  |  | 0 | ( | 0 | ) |  | 0 | ( | 0 | ) |  |  |
| Month (Epi-Week) of first vaccination |  |  |  |  |  |  |  |  |  |  | <.001 |  |  |  |  |  |  |  |  |  |  |  | 0.080 |
| December (51-53) | 91 | ( | 90 | ) |  | 19 | ( | 50 | ) |  |  |  | 18 | ( | 46 | ) |  | 1 | ( | 9 | ) |  |  |
| January (1-4) | 9 | ( | 9 | ) |  | 19 | ( | 50 | ) |  |  |  | 18 | ( | 46 | ) |  | 7 | ( | 64 | ) |  |  |
| February (5-8) | 1 | ( | 1 | ) |  | 0 | ( | 0 | ) |  |  |  | 3 | ( | 8 | ) |  | 2 | ( | 18 | ) |  |  |
| March (9-13) | 0 | ( | 0 | ) |  | 0 | ( | 0 | ) |  |  |  | 0 | ( | 0 | ) |  | 0 | ( | 0 | ) |  |  |
| April (14 - 15 ) | 0 | ( | 0 | ) |  | 0 | ( | 0 | ) |  |  |  | 0 | ( | 0 | ) |  | 0 | ( | 0 | ) |  |  |
| Unknown/not vaccinated | 0 | ( | 0 | ) |  | 0 | ( | 0 | ) |  |  |  | 0 | ( | 0 | ) |  | 1 | ( | 9 | ) |  |  |
| Analgesic intake prior to dose 1 |  |  |  |  |  |  |  |  |  |  | 0.730 |  |  |  |  |  |  |  |  |  |  |  | 0.623 |
| No | 89 | ( | 88 | ) |  | 32 | ( | 84 | ) |  |  |  | 30 | ( | 77 | ) |  | 6 | ( | 55 | ) |  |  |
| Yes | 10 | ( | 10 | ) |  | 2 | ( | 5 | ) |  |  |  | 6 | ( | 15 | ) |  | 2 | ( | 18 | ) |  |  |
| Missing | 2 | ( | 2 | ) |  | 4 | ( | 11 | ) |  |  |  | 3 | ( | 8 | ) |  | 3 | ( | 27 | ) |  |  |
| Analgesic intake after dose 1 |  |  |  |  |  |  |  |  |  |  | 0.823 |  |  |  |  |  |  |  |  |  |  |  | 0.698 |
| No | 73 | ( | 72 | ) |  | 24 | ( | 63 | ) |  |  |  | 19 | ( | 49 | ) |  | 3 | ( | 27 | ) |  |  |
| Yes | 26 | ( | 26 | ) |  | 10 | ( | 26 | ) |  |  |  | 17 | ( | 44 | ) |  | 5 | ( | 45 | ) |  |  |
| Missing | 2 | ( | 2 | ) |  | 4 | ( | 11 | ) |  |  |  | 3 | ( | 8 | ) |  | 3 | ( | 27 | ) |  |  |
| Analgesic intake prior to dose 2 |  |  |  |  |  |  |  |  |  |  | 0.437 |  |  |  |  |  |  |  |  |  |  |  | 1.000 |
| No | 84 | ( | 83 | ) |  | 27 | ( | 71 | ) |  |  |  | 25 | ( | 64 | ) |  | 6 | ( | 55 | ) |  |  |
| Yes | 15 | ( | 15 | ) |  | 7 | ( | 18 | ) |  |  |  | 11 | ( | 28 | ) |  | 2 | ( | 18 | ) |  |  |
| Missing | 2 | ( | 2 | ) |  | 4 | ( | 11 | ) |  |  |  | 3 | ( | 8 | ) |  | 3 | ( | 27 | ) |  |  |
| Analgesic intake after dose 2 |  |  |  |  |  |  |  |  |  |  | 0.322 |  |  |  |  |  |  |  |  |  |  |  | 0.126 |
| No | 57 | ( | 56 | ) |  | 16 | ( | 42 | ) |  |  |  | 21 | ( | 54 | ) |  | 2 | ( | 18 | ) |  |  |
| Yes | 42 | ( | 42 | ) |  | 18 | ( | 47 | ) |  |  |  | 15 | ( | 38 | ) |  | 6 | ( | 55 | ) |  |  |
| Missing | 2 | ( | 2 | ) |  | 4 | ( | 11 | ) |  |  |  | 3 | ( | 8 | ) |  | 3 | ( | 27 | ) |  |  |
| Abbreviations: Interquartile range (IQR), Healthcare personnel (HCP), First responders (FR), Essential and frontline workers (EFW), COVID-19-like illness (CLI); Messenger RNA (mRNA); Personal protective equipment (PPE) | | | | | | | | | |  |  |  |  |  |  |  |  |  |  |  |  |  |  |
| ¶ Occupation categories: Primary HCP (physicians, physician assistants, nurse practitioners, dentists), Other allied HCP (nurses, therapists, technicians, medical assistants, orderlies and all others providing clinical support in inpatient or outpatient settings), first responders (FR; firefighters, law enforcement, corrections, emergency medical technicians), essential and frontline workers (EFW; workers in hospitality, delivery, and retail; teachers; all other occupations that require contact within 3 feet of the public, customers, or co-workers as a routine part of their job) | | | | | | | | | |  |  |  |  |  |  |  |  |  |  |  |  |  |  |
| ‖‖ CLI was defined as reporting >1 of the following symptoms: fever, chills, cough, shortness of breath, sore throat, diarrhea, muscle or body aches, or change of smell or taste. Participants reporting non-CLI symptoms that started within 7 days prior to the collection date of the positive sample were considered to have "no CLI, but other symptoms." Participants who reported no symptoms during surveillance nor upon inquiry following SARS-CoV-2 detection were considered to have "asymptomatic +/- 14-days of PCR-detection from weekly nasal swab" | | | | | | | | | |  |  |  |  |  |  |  |  |  |  |  |  |  |  |
